# Genome-wide association study and genomic risk prediction of age-related macular degeneration in Israel

**DOI:** 10.1101/2023.09.06.23295126

**Authors:** Michelle Grunin, Daria Triffon, Gala Beykin, Elior Rahmani, Regev Schweiger, Liran Tiosano, Samer Khateb, Shira Hagbi-Levi, Batya Rinsky, Refael Munitz, Thomas W Winkler, Iris M Heid, Eran Halperin, Shai Carmi, Itay Chowers

## Abstract

**Purpose:** The risk of developing age-related macular degeneration(AMD) is influenced by genetic background. In 2016, International AMD Genomics Consortium(IAMDGC) identified 52 risk variants in 34 loci, and a polygenic risk score(PRS) based on these variants was associated with AMD. The Israeli population has a unique genetic composition: Ashkenazi Jewish(AJ), Jewish non-Ashkenazi, and Arab sub-populations. We aimed to perform a genome-wide association study(GWAS) for AMD in Israel, and to evaluate PRSs for AMD.

**Methods:** For our discovery set, we recruited 403 AMD patients and 256 controls at Hadassah Medical Center. We genotyped all individuals via custom exome chip. We imputed non-typed variants using cosmopolitan and AJ reference panels. We recruited additional 155 cases and 69 controls for validation. To evaluate predictive power of PRSs for AMD, we used IAMDGC summary statistics excluding our study and developed PRSs via either clumping/thresholding or LDpred2.

**Results:** In our discovery set, 31/34 loci previously reported by the IAMDGC were AMD associated with P<0.05. Of those, all effects were directionally consistent with the IAMDGC and 11 loci had a p-value under Bonferroni-corrected threshold(0.05/34=0.0015). At a threshold of 5x10^-5^, we discovered four suggestive associations in *FAM189A1*, *IGDCC4*, *C7orf50*, and *CNTNAP4*. However, only the *FAM189A1* variant was AMD associated in the replication cohort after Bonferroni-correction. A prediction model including LDpred2-based PRS and other covariates had an AUC of 0.82(95%CI:0.79-0.85) and performed better than a covariates-only model(P=5.1x10^-9^).

**Conclusions:** Previously reported AMD-associated loci were nominally associated with AMD in Israel. A PRS developed based on a large international study is predictive in Israeli populations.

## Introduction

Age-related macular degeneration (AMD) is the leading cause of blindness in the elderly population. The risk for developing AMD is strongly associated with the genetic background of the individual ^1,2^. In 2005, AMD was the first disease for which genome-wide association studies (GWASs) have identified risk variants ^3,4^. Via a seminal paper published in 2016, the International Age-Related Macular Degeneration Genomics Consortium (IAMDGC) has reported the genotyping of more than 30,000 AMD patients and controls of European ancestry and the discovery of 52 risk variants across 34 loci ^2^.

Israel is home to a number of populations of distinct genetic ancestry, including Ashkenazi Jews, non-Ashkenazi Jews – predominantly North-African Jews and Middle-Eastern Jews, and Arabs – predominantly Palestinians, Bedouins, and Druze. These populations are genetically diverse, having genetic ancestry related to the Middle East, Africa, and Europe, with variable mixture proportions ^5–8^. Some of the populations have experienced recent population-specific genetic drift due to founder events and endogamy ^7,9,10^ The unique genetic background of the Israeli populations suggests that the genetic architecture of AMD might be different in these populations compared to Europeans. In addition, the Israeli populations that have experienced strong genetic drift may harbor deleterious risk variants at a considerable frequency. This will increase power for discovering novel risk variants ^11^ as previously observed for other retinal diseases ^12–14^.

Previous studies of the genetic basis of AMD in Israel found that the most prominent risk variants – the genes *CFH* ^15^ and *HTRA1/ARMS2* ^16^ – were associated with AMD. However, the *C2* locus, one of the top risk loci worldwide, was not associated with AMD in Israel ^17^. The 2016 study of the IAMDGC included an Israeli cohort. However, it was analyzed jointly with the other studies, which was uninformative about Israeli-specific genetic architecture and risk variants. Searching for population-specific risk variants is important even beyond the population under study, as any discovered variants and biological pathways may provide insight into the pathogenesis of the disease.

Polygenic risk scores (PRSs) were recently developed for numerous diseases based on the results of large-scale GWASs ^18^. A PRS is the count of risk alleles carried by an individual, where each allele is weighted by its effect size (usually the log odds-ratio), as estimated by GWAS. While PRSs cannot unambiguously distinguish healthy and affected individuals (due to the small proportion of variance in disease liability they explain), individuals at the top PRS quantiles are at a particularly high risk ^19,20^. These individuals can then be subjected to personalized screening or prevention.

A number of recent papers have developed or examined PRSs for AMD, showing that the PRS has considerable power to predict disease status and disease progression ^2,21,22^. However, it is known that PRS accuracy can substantially decrease when evaluated in populations or ancestries other than the ones used for the original GWAS (usually European populations and ancestries) ^23,24^. So far, no study has examined the accuracy of an AMD PRS in any of the Israeli sub-populations, which forms a barrier to the implementation of DNA-based risk stratification.

In this paper, we used data on 558 AMD cases and 325 controls to investigate the genetic basis of AMD in the Israeli populations. Our study had three main goals. (1) To determine whether previously identified risk variants (from the IAMDGC 2016 GWAS) are associated with AMD in Israel, either across all Israeli sub-populations or in a population-specific manner. (2) To discover putative new AMD risk variants by running a GWAS in the Israeli study, anticipating that despite the small sample size, we may be able to identify risk variants that have drifted to high frequencies in the Israeli founder populations. (3) To evaluate the accuracy in the Israeli population of a PRS generated based on the IAMDGC GWAS. We show that the vast majority of previously discovered risk variants are also associated with AMD in Israel, Accordingly, a PRS based on previously discovered variants has high predictive power. While our study was too small for discovering new risk variants at a genome-wide significance level, our study suggested a number of putative associations at an attenuated significance threshold.

## Results

### Replication of known AMD loci

A previous large-scale AMD GWAS by the IAMDGC (n=33,976 ^2^) has discovered 34 associated loci. We examined the association of these loci with AMD status in our Israeli discovery set (403 AMD cases and 256 controls). Using the SNP with the lowest p-value in each locus, we found that most loci (31/34) were associated with AMD at a nominal significance level of P<0.05 with a direction of effect consistent with that of the IAMDGC (Supplementary Tables 2 and 3). The number of loci associated at the Bonferroni correction threshold (0.05/34=0.0015) was 11/34 (Supplementary Table 2). The top ranked loci were *CFH* (P=1.6·10^-9^) and nearby loci on chr1, and *ARMS2*/*HTRA1* (P=3.4·10^-9^, 5.1·10^-9^, respectively). The next significant locus was near *SYN3* (P=5.7·10^-5^). We note that replication was to some extent expected, given that the majority of the Israeli cohort was included in the IAMDGC. Association statistics for the known AMD risk loci for AJ (242 cases and 136 controls) and Arabs (36 cases and 30 controls) are reported in Supplementary Tables 4 and 5.

### Discovery GWAS

We next ran a GWAS in our discovery set (AMD cases: n=403, controls: n=256). No novel variant was associated at the genome-wide significance threshold of 5·10^-8^. Setting a more liberal threshold of 5·10^-5^, and excluding variants in known risk loci, we identified four suggestive associations in the genes *C7orf50*, *IGDCC4*, *FAM189A1*, and *CNTNAP4* (Table 1; Figure S2). None of these SNPs were associated with AMD in the IAMDGC data (P≥0.04, Table 1). The variant rs116928937 in *IGDCC4* is exonic. Its allele frequency in European Americans was 1.23% (In the exome variant server), compared to 2.66% here. It is a missense variant (c.3188G>T), and according to Polyphen ^25^ it is “probably-damaging”.

**Table 1.** Statistics for the association of SNPs with AMD status in our Israeli discovery cohort. Genomic coordinates are in hg19. The gene is the nearest to the SNP. Allele frequencies were computed in gnomAD, for either non-Finnish Europeans (NFE) or Ashkenazi Jews (AJ). The first four rows provide details on four SNPs associated with AMD in our discovery cohort with p-value <5·10^-5^. The last row presents details on the top-associated SNP in the AJ subset of our cohort.

| SNP ID | Genomic Position (hg19) | Gene | Annotation | P-value (entire cohort) | Odds ratio (95% confidence interval) Entire cohort, AJ subset | Minor allele frequency (cases/controls) | Minor allele frequency (gnomAD NFE/AJ) | P-value in IAMDGC - leave one out | Replication P-value Genotype (Fisher's exact) |
| --- | --- | --- | --- | --- | --- | --- | --- | --- | --- |
| rs12701455 | chr7:1055409 | C7orf50 | | $5.05 \times 10^{-6}$ | 0.5 (0.26-94) | 0.147/0.251 | 0.309/0.212 | 0.78 | 0.028 |
| rs116928937 | chr15:65677446 | IGDCC4 | Missense | $3.73 \times 10^{-6}$ | 0.13(0.02-0.83), 0.07 | 0.008/0.055 | 0.0185/0.0064 | 0.89 | 1 |
| rs1506825 | chr16:76483019 | CNTNAP4 | | $6.91 \times 10^{-6}$ | 0.57(0.34-0.96) | 0.4373/0.5703 | 0.455/0.469 | 0.04 | 0.85 |
| rs1195500 | chr15:29687047 | FAM189A1 | | $2.265 \times 10^{-5}$ | 0.51(0.27-0.96), 0.37 | 0.129/0.227 | 0.26/0.15 | 0.27 | 0.00001 |
| rs11689931 (AJ specific) | chr2:206440979 | PARD3B | | $3.19 \times 10^{-6}$ for AJ | 0.07 | 0.20/0.37 in AJ | 0.301/0.306 | | Was not tested |

We attempted to replicate the association of these four loci in a replication set of n=155 AMD cases and n=69 controls (Supplementary Table 1). We applied a Bonferroni corrected threshold of 0.05/4=0.0125. Only the SNP in *FAM189A1* (rs1195500, chr15:29687047) replicated in the Israeli population (P<0.0001 in Fisher’s exact test in both genotype and allele testing). The SNP rs12701455 (*C7orf50*) attained a p-value of 0.029 in the genotype-based test (Supplementary Table 1).

### Evaluating a polygenic risk score for AMD

We developed polygenic risk scores (PRSs) for AMD in Israel based on the results of the IAMDGC GWAS and using two methods. The first method is clumping and thresholding (C+T), in which the most strongly associated SNP is retained from each LD block, as long as its p-value is under a threshold. The second method is LDpred2, which accounts for the influence of LD on effect sizes and incorporates a non-zero prior probability for having null effects. We generated nine C+T PRSs, corresponding to different p-value thresholds (exponentially increasing between 5·10^-8^ and 1), and four LDpred2 PRSs, corresponding to different values of the proportion of SNPs with non-zero effects and a sparsity parameter. For each PRS, we used logistic regression to predict AMD status based on age, sex, the first two principal components (a proxy of ancestry), and the PRS. We also fit a logistic regression model with covariates only. We used 5-fold cross-validation to evaluate the accuracy of the various models, which we quantified using AUC (the area under the receiver operator curve (ROC)).

We compare the ROCs of the top C+T model, the top LDpred2 model, and the covariates-only model in Figure 1. For C+T, the AUC was highest (0.79; 95% confidence interval (CI): 0.75-0.82) for the most stringent p-value threshold (5·10^-8^), for a PRS that included 360 variants. Interestingly, the AUC decreased monotonically with increasing p-value thresholds (Figure S3). The top LDpred2 model (parameters *p*=0.056 and sparsity on) had a slightly higher AUC (0.82; 95% CI: 0.79-0.85) than the top C+T model. The covariates-only model had a significantly lower AUC (0.72; 95% CI: 0.69-0.76). This was also confirmed by DeLong’s test for two correlated ROC curves (P=5.1·10^-9^). Overall, our results suggest that including the PRS in the prediction model improves accuracy.

**Figure 1.**
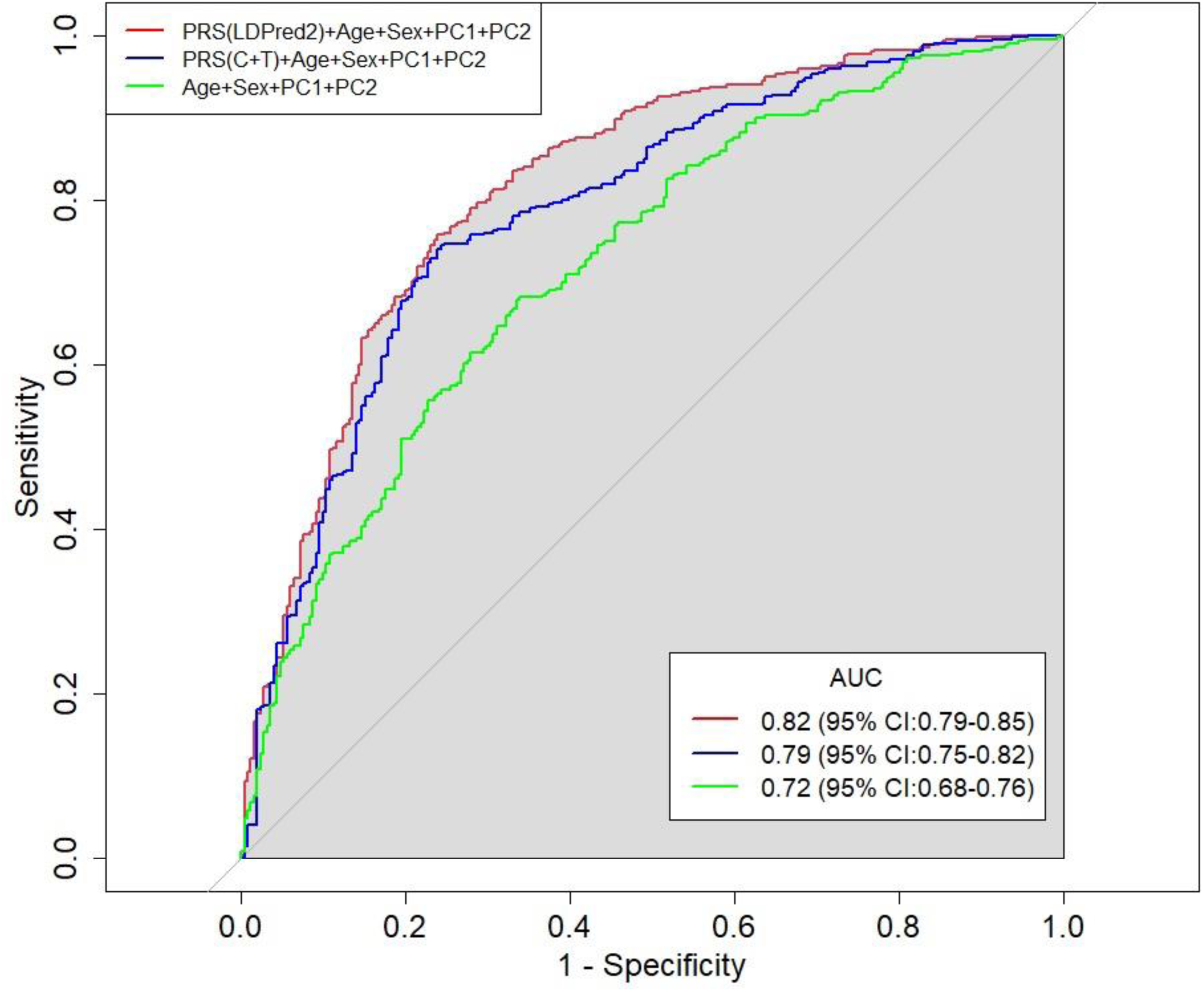
Prediction accuracy of selected PRS models. We show the ROC curve for the following AMD prediction models: the top C+T PRS (Figure S3) + covariates (age, sex, PC1, and PC2); the top LDpred2 PRS + covariates; and a covariates-only model. The C+T PRS parameters were r²>0.5 and P<5·10^-8^, and the LDpred2 PRS parameters were *p*=0.056 and sparse=TRUE. The AUC estimates (after cross-validation) are indicated on top of the plot.

We show the distribution of the top LDpred2 PRS in cases and controls in Figure 2A. The PRS distribution is different between cases and controls; however, considerable overlap exists. In Figure 2B, we plot the proportion of cases in each quintile of the LDpred2 PRS, demonstrating that the proportion of cases steadily increases with increasing quintiles. Finally, we used Spearman’s correlation test to assess the correlation between age at diagnosis (measured here as age at blood draw) and the PRS among AMD cases. In Figure S4, we show a modest, yet significant, negative correlation between the variables (ρ=-0.18, p-value=0.0003, using the best LDpred2 PRS), suggesting that the PRS may be associated not only with disease status but also with age of onset.

**Figure 2.**
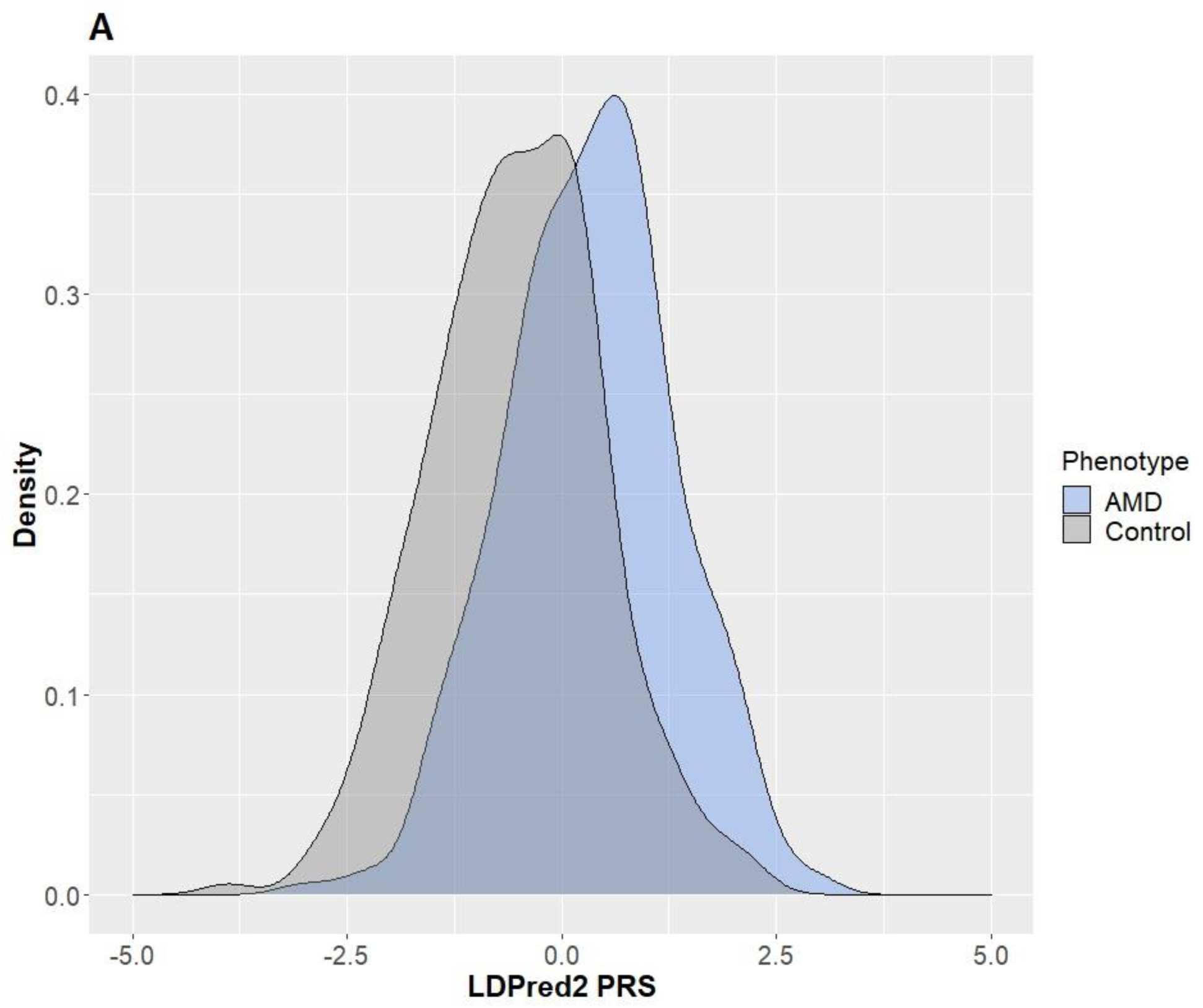
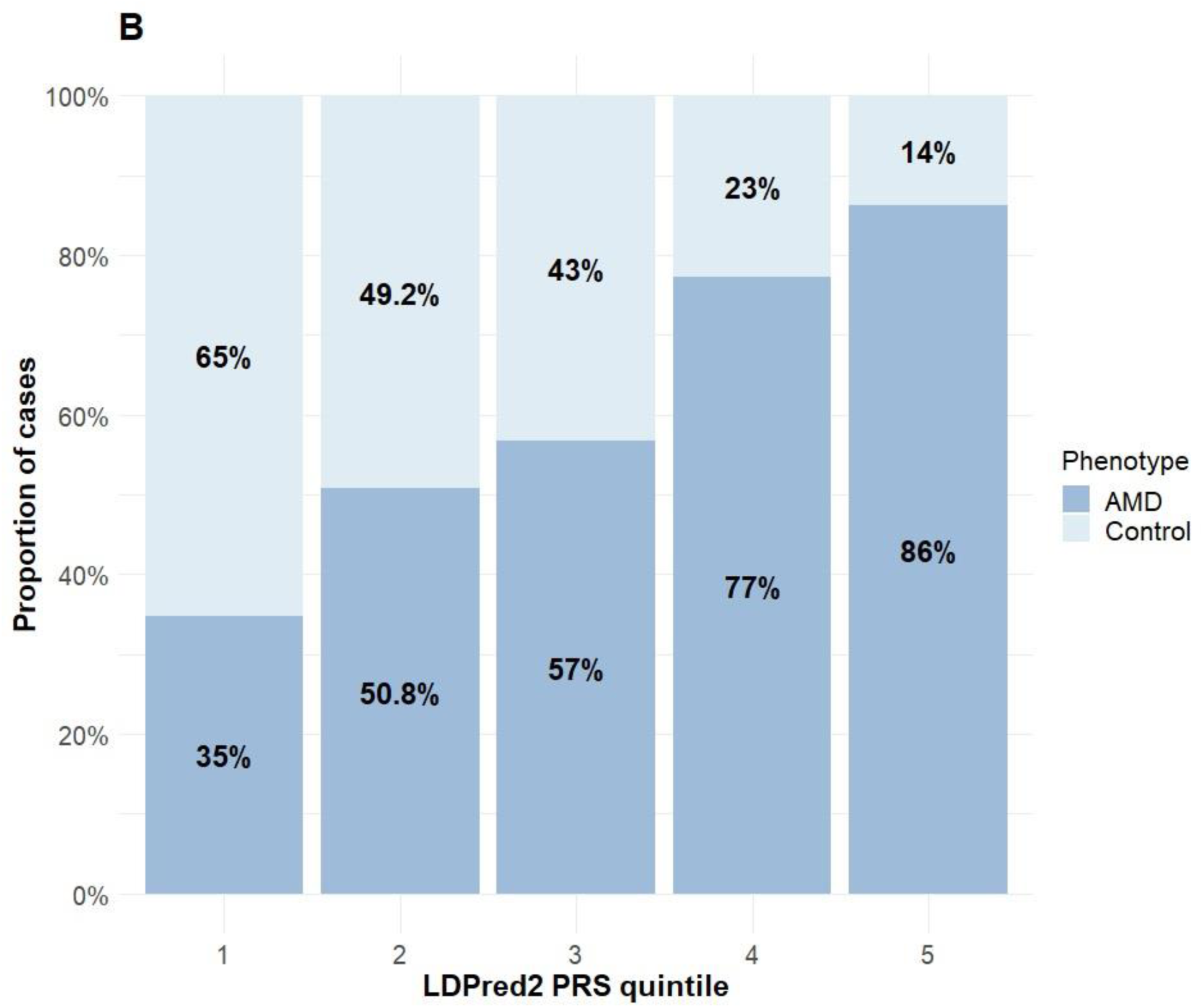
Comparing the PRS between cases and controls. (A) The density of the top LDpred2 PRS (after regressing out the first two principal components) in AMD cases and controls in our study (n=403 and 259, respectively). (B) The proportion of AMD cases in our study by PRS quintiles. We again used the top LDpred2 PRS.

## Discussion

In this work, we studied the genetic basis of AMD in the Israeli populations. We confirmed that most of the known risk loci for AMD, as previously identified in a large international study, are also associated with AMD in Israel. This suggests that the genetic architecture of AMD is similar between the Israeli and other populations. We then performed a genome-wide association study in our cohort in an attempt to identify novel risk variants. As expected due to the small size of our cohort, no novel variants were detected at the genome-wide significance threshold. Setting a more relaxed threshold of 5·10^-5^, we identified four suggestive variants. One of these variants (rs1195500, in *FAM189A1*) replicated, after Bonferroni correction, in a small second set of cases and controls.

The evaluation of the AMD PRS in our Israeli cohort suggested multiple conclusions. First, the LDpred2 PRS had relatively high accuracy (AUC=0.82), significantly better compared to not including the PRSs (Figure 1). Second, with the simple clumping and thresholding approach, accuracy increased as more stringent p-value thresholds were used (Figure S3). This could indicate that AMD is not as polygenic as other diseases. Third, as expected^26,27^, LDpred2 performed better than the C+T approach (Figure 2A). Finally, high AMD PRS in our study associated not only with disease risk, but also with a lower age of onset (Figure S4), as seen for diseases in other domains^28,29^. Prospective studies will be required to further validate this finding.

Our results for high predictive power of the AMD PRS are in line with previous studies ^30,31^ in individuals of European ancestry. The transferability of the PRS to the Israeli population is perhaps expected given that the majority of our subjects had Ashkenazi Jewish ancestry, and given that PRSs for other diseases and traits were shown to have high accuracy in Ashkenazi Jews ^32–35^. The transferability of PRSs into Ashkenazi Jews may be due to the high percentage of European ancestry in this population ^7^. It is also consistent with our replication of the known risk loci. Our sample size was too small to evaluate the PRS accuracy in other sub-populations, which could be the goal of future studies. Further improvement of the PRS may be achieved via denser genotyping or larger and more diverse imputation reference panels. Additionally, multiple methods can leverage even small samples from a target non-European population to improve a PRS constructed using large GWASs in Europeans ^36,37^. However, such efforts will require additional samples for evaluation of the resulting PRSs.

## Materials and Methods

Our discovery set consisted of 403 AMD cases and 256 controls (659 total) recruited at Hadassah Medical Center, as previously reported ^38^. Our cases included both atrophic and neovascular (a more advanced) AMD. The subjects’ mean age was 75.4 years (SD: 2.76, range: 60-97) and 44.6% were female. The criteria for inclusion of AMD patients were: age >60, AMD diagnosis according to AREDS (Age-Related Eye Disease Study) ^39^, and choroidal neovascularization (CNV) and/or geographic atrophy. Diagnosis was also determined according to fluorescein angiogram and optical coherence tomography. Participants were included in all stages of AMD. We excluded individuals with other retinal diseases and individuals with other potential CNV causes such as myopia, trauma, or uveitis. Controls were over the age of 60 with a normal fundus examination and similar systemic exclusion criteria. The study was approved by the institutional ethics committee. All subjects signed informed consent forms that adhered to the tenets of the declaration of Helsinki.

We genotyped all subjects on the custom chip that was developed for the IAMDGC. Genotyping on this chip was performed either via the IAMDGC (at the Center for Inherited Disease Research (Johns Hopkins, USA)) or at the genomics core facility of the Technion (Israel), as previously described ^31^. The custom chip, which was previously described, contains ≈250,000 tagging markers for imputation and ≈250,000 custom markers for AMD ^2^.

We imputed the genomes of our subjects with the following reference panels: the 1000 Genomes Project (n=2504) ^40^ and the Ashkenazi Genome Consortium (n=128) ^6^. This strategy was shown to have the highest accuracy for imputing Ashkenazi genomes ^41^ and was applied here, given that 60% of our subjects have Ashkenazi ancestry. Unfortunately, a reference panel for non-Ashkenazi Jews or for the non-Jewish populations of Israel does not yet exist. We phased our genomes using SHAPEIT ^42,43^ and performed imputation using a standard protocol ^43,44^. We describe next the post-imputation quality control (QC) pipeline, as we previously developed ^38,45^.

The chip was imputed to 37,126,112 variants. We performed QC according to standard protocols to remove low-quality variants and samples ^46^. We excluded variants with imputation quality score R^2^<0.6, variants with minor allele frequency <0.01, and variants in Hardy-Weinberg disequilibrium (PLINK 1.9 ^47,48^). The sex of patients was confirmed using the sex-check option in PLINK. We excluded individuals who were related, having PIHAT>0.3 in PLINK. We performed principal components analysis (PCA) in PLINK and GCTA ^49^ to account for population stratification; the first two principal components were used as covariates in the association analysis (Figure S1). The final variant count after filtering was 5,353,842 variants in 403 AMD patients and 256 controls.

We performed the discovery GWAS on case-control status using logistic regression in PLINK. To account for population stratification, we used the first two principal components as covariates. The other covariates were age at blood draw and sex. We generated Manhattan and Q-Q plots with *qqman*. For genome-wide significance, we used a p-value threshold of 5·10^-8^. To detect suggestive associations, we used a threshold of 5·10^-5^. We computed the frequency of risk alleles (either in Europeans or in Ashkenazi Jews) using gnomAD (http://gnomad.broadinstitute.org/) and, if exonic, also in the Exome Variant server (http://evs.gs.washington.edu/EVS). Variants that were outside gene boundaries were reported to nearest gene. Variants contained within a gene were reported with that gene.

To determine whether previously discovered associations replicate in our study, we considered variants within the 34 known loci that were identified in the IAMDGC 2016 GWAS ^2^ (Table 5 in the IAMDGC GWAS paper). For each locus (LD block) we retained the variant with the lowest p-value. We considered a nominal significance level of P=0.05 or a Bonferroni corrected level of P=0.05/34=0.0015.

To test for population-specific replication, we separately studied Ashkenazi Jews (AJ; 242 cases, 136 controls) and Arabs (36 cases and 30 controls). We identified AJ by self-report, requiring both parents to have AJ ancestry, and via their clustering in a principal components analysis with the Ashkenazi reference genomes (Figure S1). We identified Arab subjects based on self-report (36 cases and 30 controls). We considered all variants in linkage disequilibrium (LD; r^2^>0.05 in AJ, using hg19 linkage blocks as per the original Fritsche et al 2016 paper) to belong to the same locus. We note that 549/649 of our subjects were part of the original IAMDGC GWAS ^2^ (out of a total of 33,976 individuals). Therefore, some degree of replication is expected just by virtue of this sample overlap. However, given that the Israeli samples were less than 2% of the total IAMDGC sample, the effect of the overlap is expected to be small.

To replicate putative discoveries from the present study, we recruited additional 155 AMD cases and 69 controls (total 224) according to the same criteria as in the original discovery set. We used this case/control sample to validate the suggestively associated variants from the discovery set. Four variants passed the 5·10^-5^ genome-wide threshold in the discovery set, after excluding variants in known AMD risk loci. We genotyped these four variants in our entire replication set using the KASP assay (LGC Group, Middlesex, UK) with custom primers. All heterozygotes were confirmed using Sanger sequencing (Macrogen, Seoul, Korea). We tested the association using EPACTS (https://genome.sph.umich.edu/wiki/EPACTS) and R using two tests. For each SNP, an allelic test compared the proportion of minor alleles between cases and controls. A genotypic test compared the proportion of homozygotes to the minor allele out of all homozygotes between cases and controls.

To generate a polygenic risk score for AMD, we first performed quality control according to standard protocols. In parallel, we excluded the Israeli samples from the IAMDGC dataset and re-ran the GWAS analysis (remaining n=33,515; the “base” study). We used the resulting effect sizes to compute the PRS for individuals in the Israeli study (n=659; the “target” study). We removed variants with strand-ambiguous variants from the base study’s summary statistics. Duplicated variants were removed from both studies, separately. Variants with mismatching alleles were also removed. This has left 4,070,992 overlapping variants between the two studies (directly genotyped or imputed).

We generated polygenic risk scores using two approaches for variant selection: clumping and thresholding (C+T) and LDpred2. Briefly, in C+T, index variants are sequentially selected based on having the lowest p-value, and nearby variants in LD with the index variants are removed. Index variants with p-value under a threshold are retained^50,51^. We computed LD (r^2^) using PLINK and the target study. We set the clumping parameters to r²>0.5 and ±500 kb and used nine p-value thresholds: 5·10^-8^, and 10^-7^, 10^-6^, 10^-5^, 10^-4^, 0.001, 0.01, 0.1, and 1. The minimum p-value cutoff was set to match the IAMDGC genome-wide significance threshold.

LDpred2 is a Bayesian method for deriving polygenic scores based on summary statistics while explicitly accounting for LD. Briefly, causal effect sizes are assumed to be a mixture of a normal distribution and a point mass at zero. Posterior mean effects are computed using Gibbs sampling based on the LD matrix and an estimate of the heritability^26,27^. We set the SNP heritability (*h*^2^) to 0.47 based on a previous IAMDGC estimate^31^ and used five proportions of causal variants (*p*) spaced on a log scale: 10^-5^, 1.8·10^-4^, 0.0032, 0.056, and 1. We also included a third parameter of sparsity (true/false). The analysis was restricted to HapMap3 variants. The LD matrix was computed using the target study. We used the R package bigsnpr to compute the LD matrix and generate the grid of scores ^27^. To avoid confounding by ancestry, in both methods we regressed the scores on the first two principal components and used the residuals as the scores in subsequent analyses.

We used PLINK to calculate PRSs for each of the 659 subjects. Overall, we obtained nine C+T PRSs (nine p-value cutoffs) and ten LDpred2 PRSs, of which four were reported as valid by LDpred2 (p=0.056, sparse=FALSE; p=0.056, sparse=TRUE; p=1, sparse=FALSE; p=1, sparse=TRUE). To evaluate the accuracy of each score, we used logistic regression of the disease status on the PRS and the following covariates: age, sex, PC1, and PC2. We also included a logistic regression model based on covariates only. We measured the accuracy of each model using the area under the curve (AUC) of the receiver operator curves (ROC), computed using 5-fold cross validation. We used the R package *pROC* (https://cran.r-project.org/web/packages/pROC/pROC.pdf) to generate and analyze ROCs, AUCs, and AUC confidence intervals (ci.auc ()), and the R package *caret* (https://cran.r-project.org/web/packages/caret/vignettes/caret.html) for cross-validation. Individuals with missing age data were excluded from the analysis (four cases and five controls).

We visually inspected the discriminatory power of the PRS using plots of the density of the PRS in cases and controls (using kernel density estimation), and the proportion of AMD cases across quintiles (fifths) of the PRS distribution. Both plots were generated with the R package *ggplot2* (https://cran.r-project.org/web/packages/ggplot2/index.html). Finally, we computed Spearman’s rank correlation coefficient to examine the association between the PRS and age at blood draw (as a proxy of the age at diagnosis) among AMD cases. Plots were generated with R package *ggpubr* (https://cran.r-project.org/web/packages/ggpubr/index.html).

## Data Availability

The full IAMDGC dataset values can be accessed at: http://amdgenetics.org/ including the entire IAMDGC and the Jerusalem dataset specifically. In addition, the GWAS summary statistics and code utilized in this manuscript can be found by contacting the corresponding author via reasonable request. The Related Manuscript Variant supplementary file contains all nomenclature for HGVS for all variants.

## Author Contributions

MG, DT, SC, IC, ER conceived and designed the work, MG, DT, ER, RS, RM designed and performed the experiments and data analysis, MG, DT, ER, RS, GB, LT, SK, SH-L, BR, RM, IC and SC provided data availability and data extraction and analysis, MG, DT, SC, IC wrote the manuscript, MG, DT, SC, IC, ER, RS, EH provided manuscript feedback, revision, and drafts.

## Ethical Approval

The study was approved by the institutional ethics committee of Hadassah Medical Center. All subjects signed informed consent forms that adhered to the tenets of the declaration of Helsinki. The IAMDGC study participants were previously ascertained by IAMDGC cohorts as described in Fritsche et al, 2016, Nature Genetics. All participants provided informed consent, and the study was approved by institutional review boards as previously described.

---

**Figure S1.**
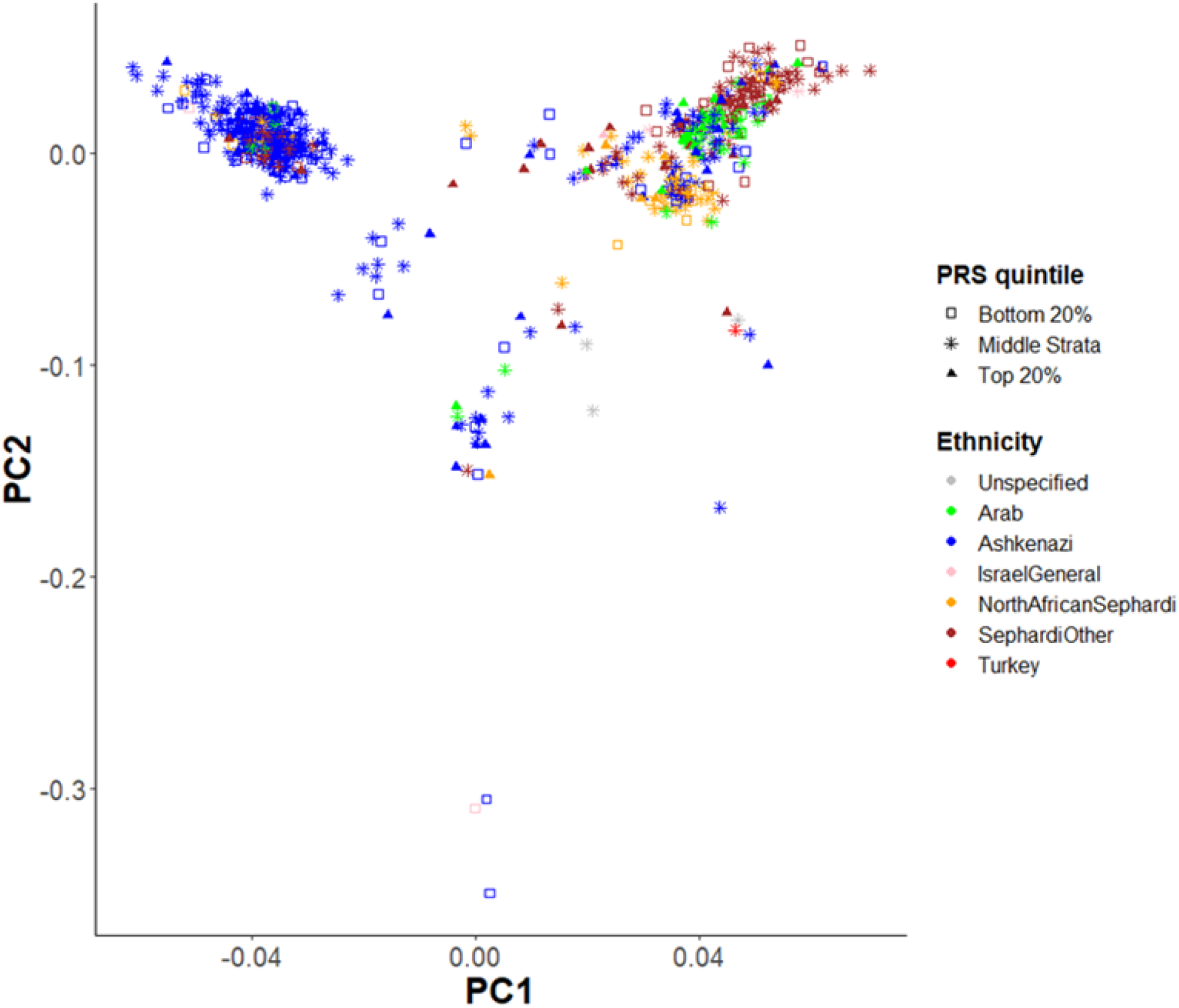
A PCA plot of the entire discovery cohort. Each symbol represents a single individual. Individuals are color coded based on their self-reported ancestry: Ashkenazi (n=378), North African Sephardi, Turkey, and other Sephardi (n=215), Arab (n=66), and Israel general (n=10). The shape of each symbol corresponds to the AMD PRS quintile (see legend).

**Figure S2.**
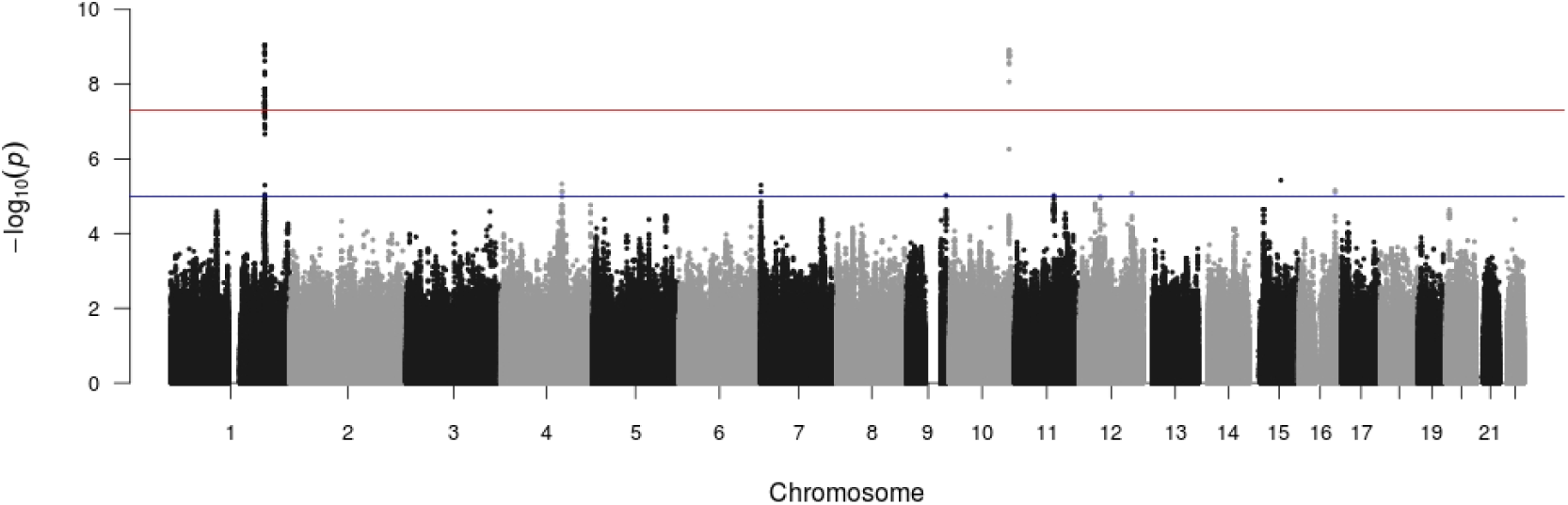
A Manhattan plot for the GWAS of AMD in the Israeli discovery cohort (n=659). The X axis indicates chromosomal position, and the Y axis indicates significance, as measured by –log_10_P.

**Figure S3.**
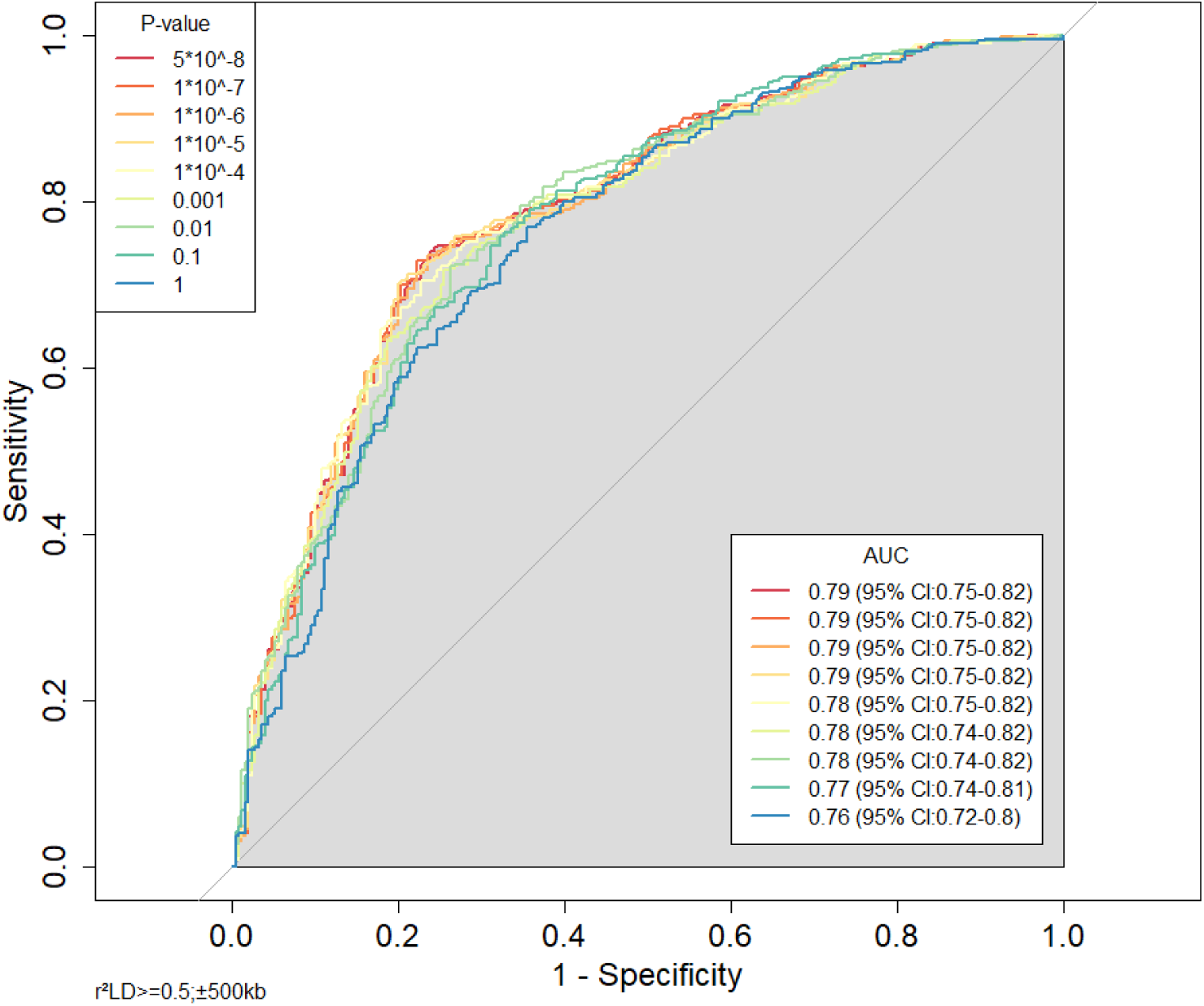
Accuracy of logistic regression models for predicting AMD disease status using clumping+thresholding (C+T) PRSs. The models also included the following covariates: age, sex, PC1, and PC2. Each curve corresponds to one of nine p-value cutoffs (see legend). The AUC values (after cross-validation) are presented in the legend. CI: confidence interval. Prediction accuracy increased as the p-value threshold decreased.

**Figure S4.**
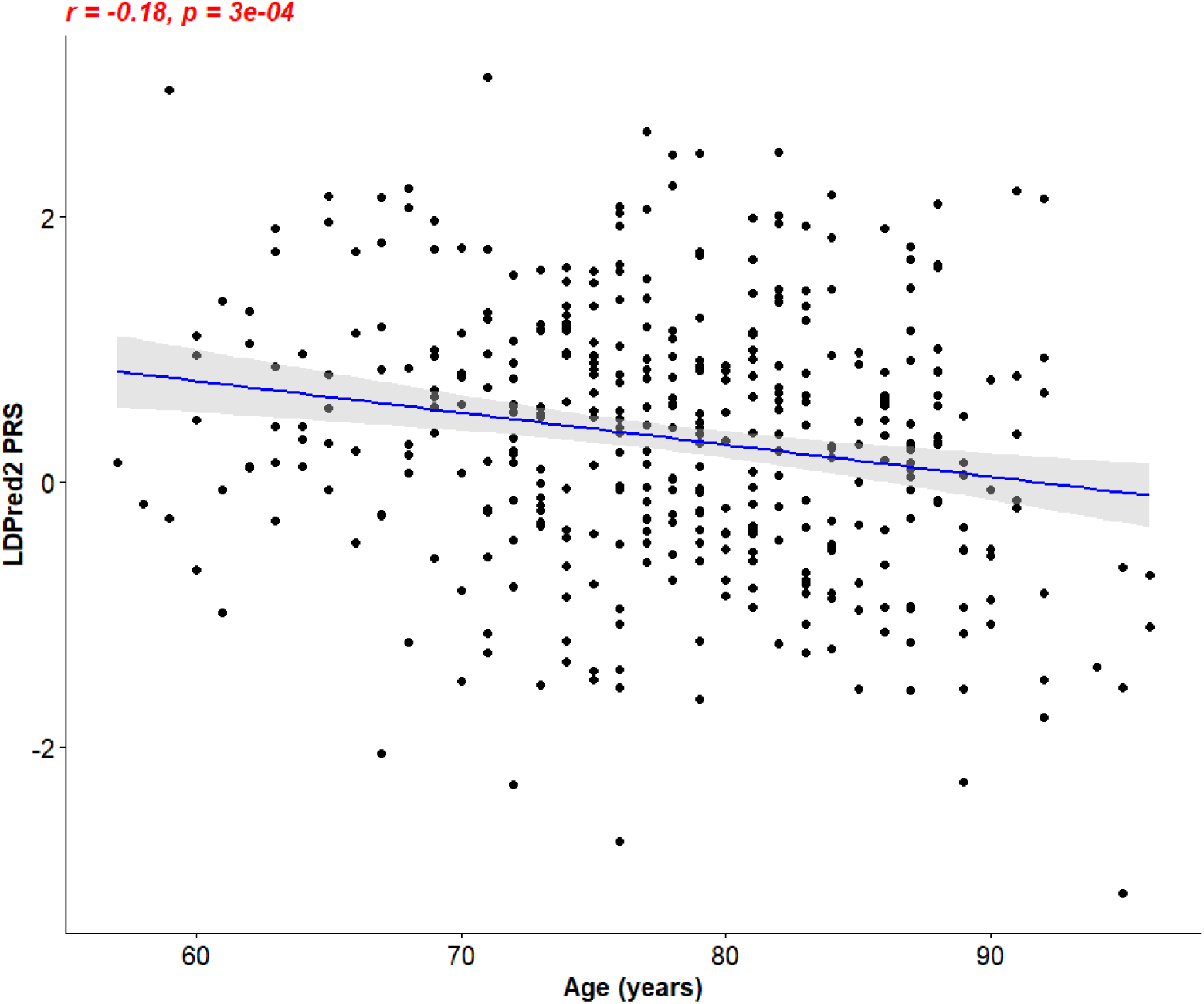
A scatter plot of age at blood draw and PRS among AMD cases (n=399). We used the best-performing LDpred2 PRS. The PRS was adjusted for PC1 and PC2 to account for confounding by ancestry. The presented age is a proxy for the age at diagnosis. The linear regression parameters are indicated (r=-0.18, P=0.0003). The regression line is also shown, along with the 95% confidence interval (gray band).

**Supplementary Table 1:**
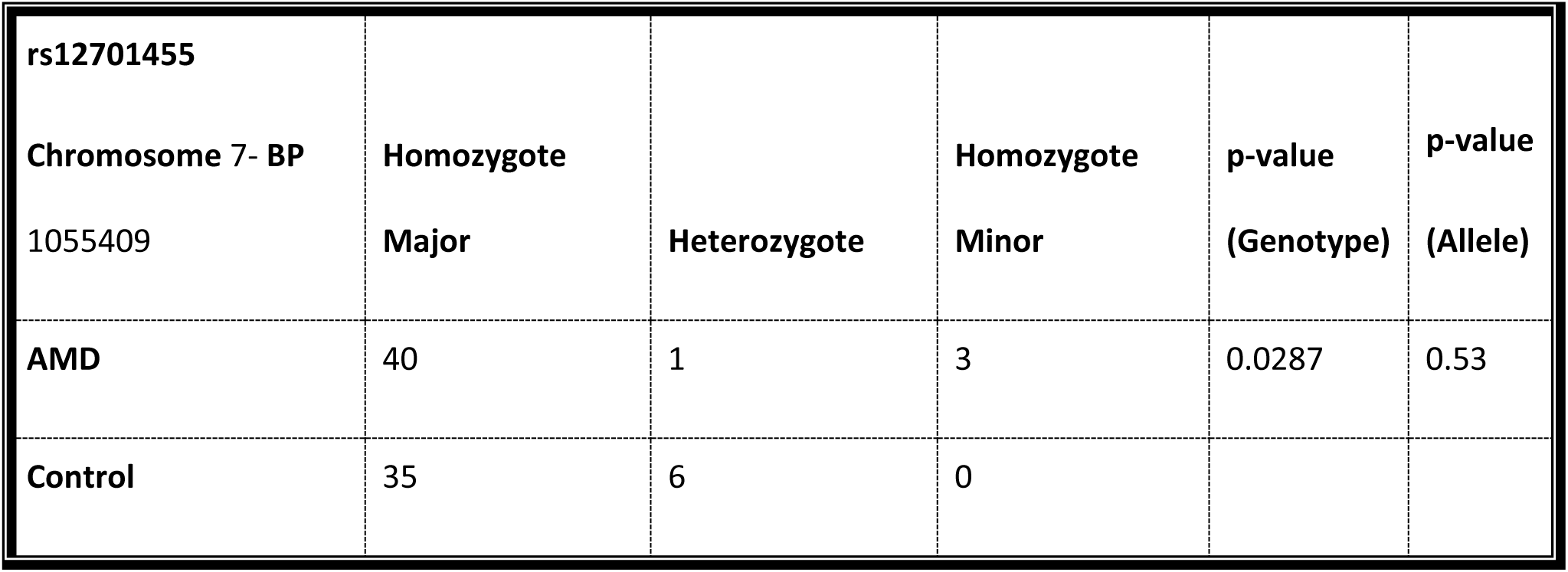
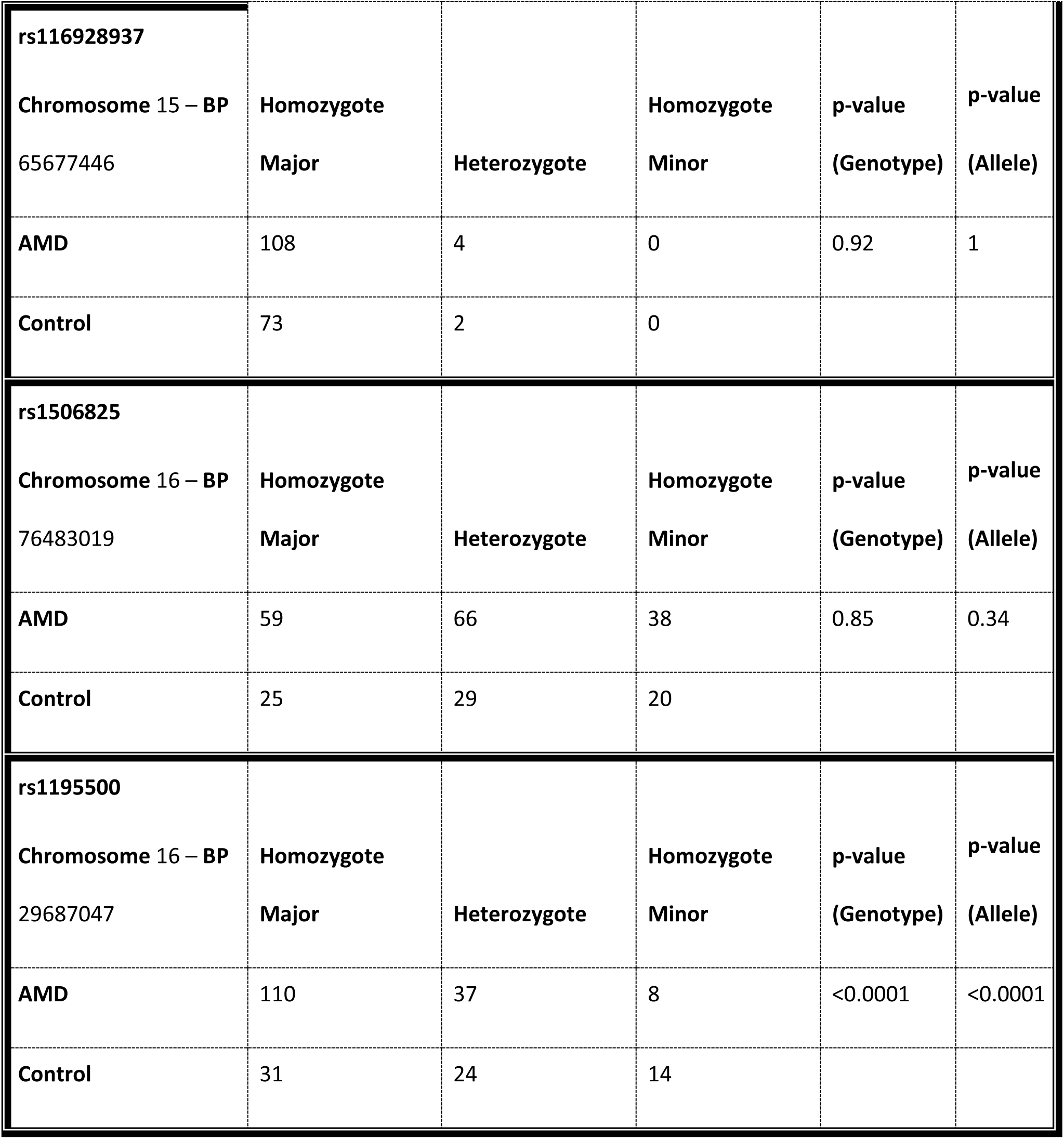
Replication cohort results (n=224) for validation of four top associated SNPs from the AMD GWAS in the Israeli population. Genotype and allelic p-values according to Fisher’s exact test are given. The bold bars separate the four SNPs tested for validation. The table provides their genotypes in the replication cohort tested and the p-values.

**Supplementary Table 2.** Association statistics of 27 variants in 11 known AMD risk loci that replicated in the Israeli discovery set after Bonferroni correction (threshold 0.05/34=0.0015). For each variant, we provide the gene, chromosome (Chr), basepair (BP), odds ratio (OR), 95 percent confidence interval (95% CI) and p-value (P). The variants are sorted by their p-value.

|  | Gene | Chr | BP | OR | 95%CI | P |
| --- | --- | --- | --- | --- | --- | --- |
| 1 | CFH | 1 | 196695161 | 0.4687 | [0.25-0.85] | 1.56E-09 |
| 2 | ARMS2 | 10 | 124214448 | 2.071 | [1.16-3.84] | 3.424E-09 |
| 3 | HTRA1 | 10 | 124221270 | 2.061 | [1.15-3.79] | 5.077E-09 |
| 4 | CFHR5 | 1 | 196978615 | 0.5161 | [0.29-0.89] | 5.195E-08 |
| 5 | CFHR2 | 1 | 196927791 | 1.894 | [1.11-3.39] | 1.163E-07 |
| 6 | CFHR4 | 1 | 196870299 | 0.436 | [0.21-0.96] | 0.00000762 |
| 7 | KCNT2 | 1 | 196406715 | 0.5845 | [0.33-0.96] | 0.00001443 |
| 8 | F13B | 1 | 197012111 | 1.7 | [1.03-2.83] | 0.00001487 |
| 9 | SYN3 | 22 | 33047598 | 0.3696 | [0.13-0.97] | 0.00005685 |
| 10 | ASPM | 1 | 197094030 | 1.616 | [1.01-2.62] | 0.0000688 |
| 11 | ZBTB41 | 1 | 197132378 | 0.4372 | [0.19-0.99] | 0.0000732 |
| 12 | PLEKHA1 | 10 | 124148167 | 0.6403 | [0.39-0.98] | 0.0001034 |
| 13 | VPS29 | 12 | 110935268 | 0.5591 | [0.31-1.01] | 0.000218 |
| 14 | CRB1 | 1 | 197199434 | 1.568 | [0.99-2.5] | 0.0002242 |
| 15 | PPTC7 | 12 | 110994068 | 0.5699 | [0.24-1.01] | 0.0002391 |
| 16 | ZNF557 | 19 | 7083629 | 1.576 | [0.98-2.51] | 0.0002624 |
| 17 | HVCN1 | 12 | 111099721 | 0.5184 | [0.26-1.03] | 0.0004554 |
| 18 | MICALL1 | 22 | 38318897 | 1.48 | [0.97-2.25] | 0.0007491 |
| 19 | SMC5 | 9 | 72920724 | 0.427 | [0.17-1.06] | 0.0008093 |
| 20 | PPP1CC | 12 | 111160003 | 0.5937 | [0.34-1.04] | 0.0008394 |
| 21 | TCTN1 | 12 | 111081197 | 0.5847 | [0.32-1.04] | 0.0008647 |
| 22 | HLA-B | 6 | 31324864 | 0.4779 | [0.22-1.06] | 0.0008669 |
| 23 | RAD9B | 12 | 110948906 | 0.5532 | [0.29-1.0504] | 0.001058 |
| 24 | EIF3L | 22 | 38273303 | 1.476 | [0.97-2.25] | 0.001161 |
| 25 | TTC23L | 5 | 34840841 | 0.6909 | [0.46-1.033] | 0.001183 |
| 26 | ACHE | 7 | 100491753 | 0.1589 | [0.02-1.18] | 0.001267 |
| 27 | KIAA0100 | 17 | 26956537 | 0.6755 | [0.44-1.04] | 0.001267 |

**Supplementary Table 3.**
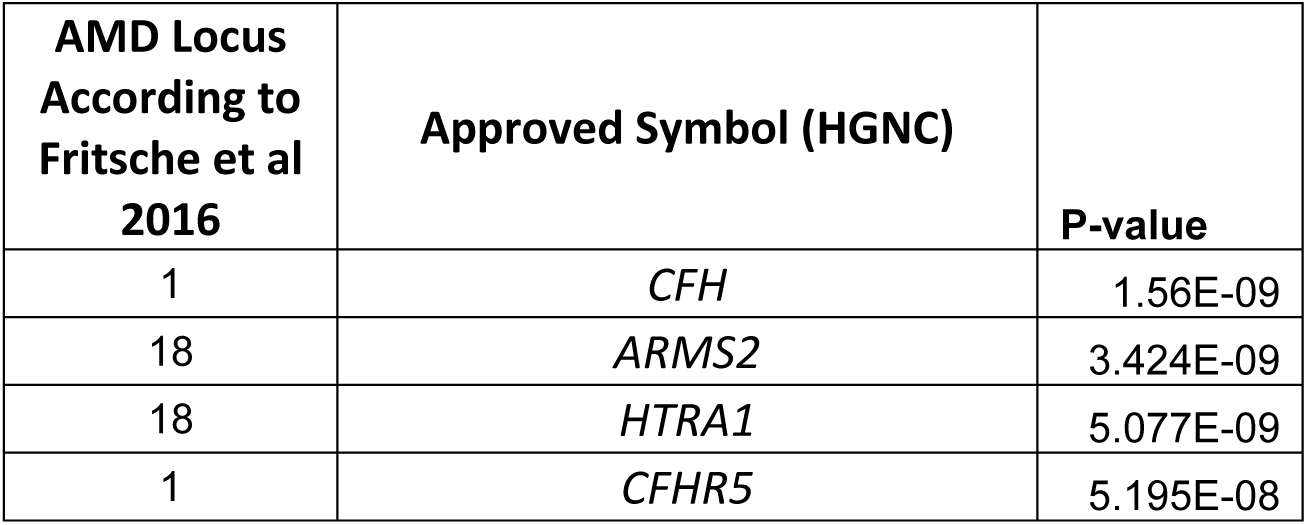
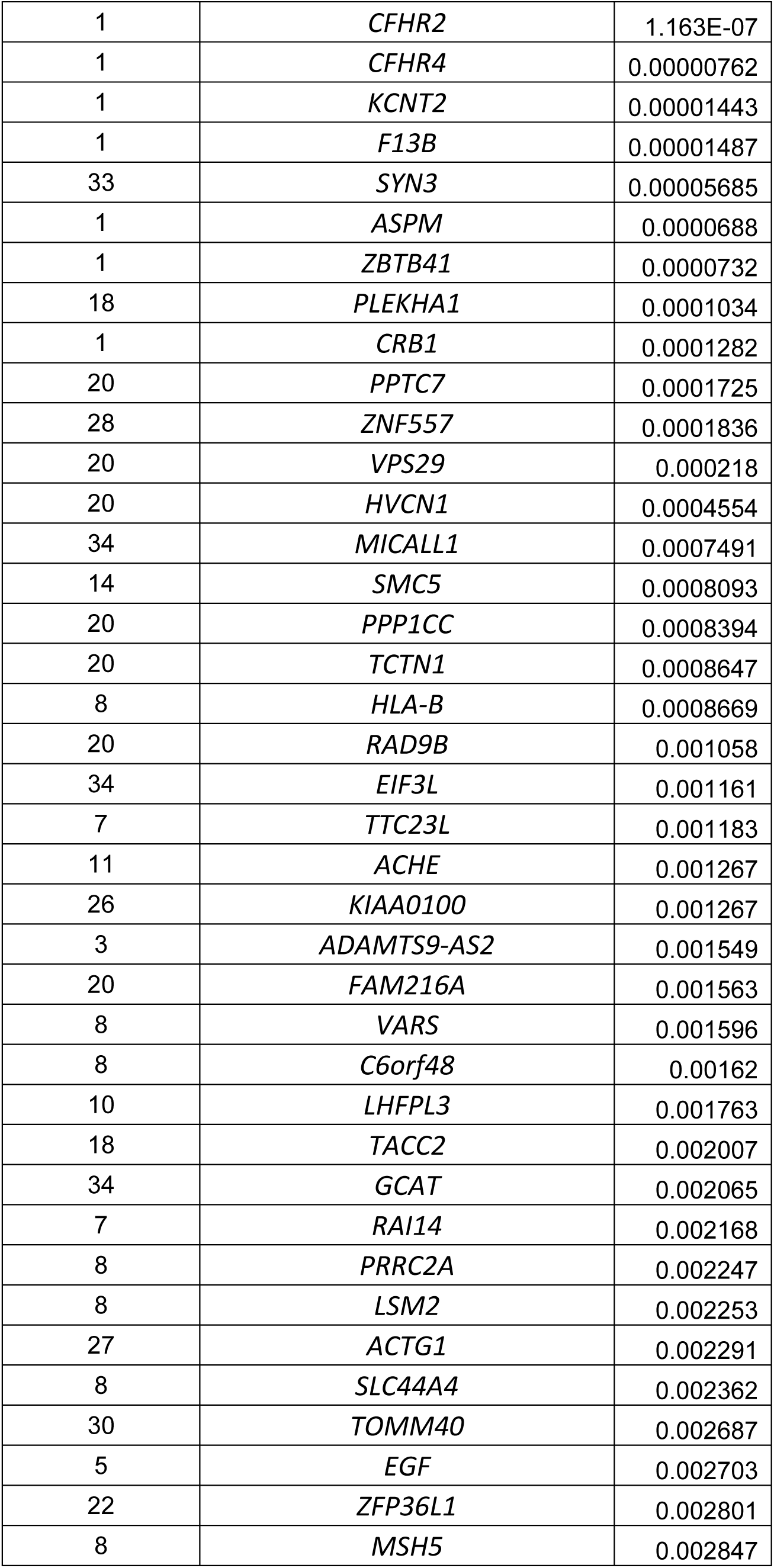
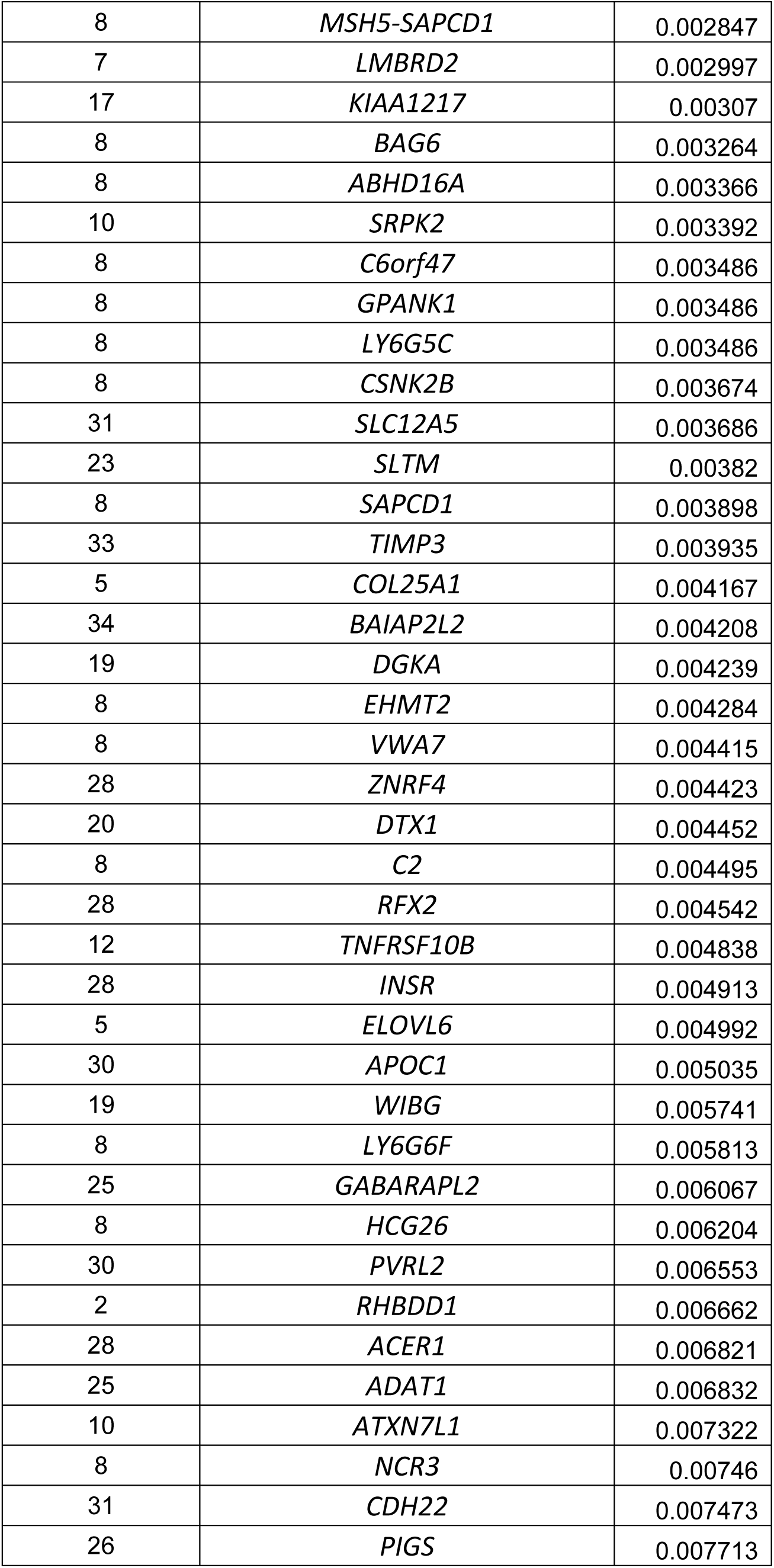
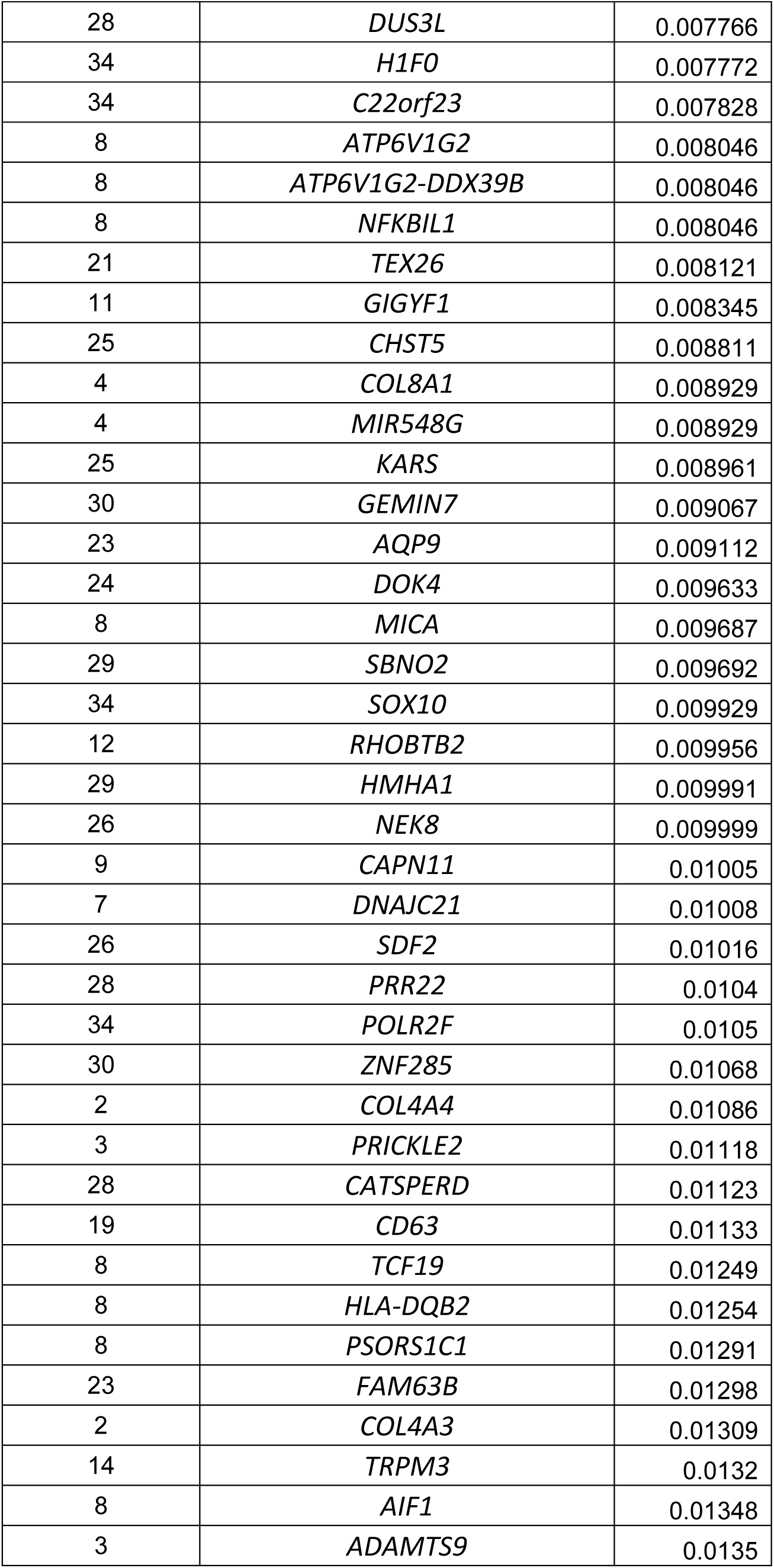
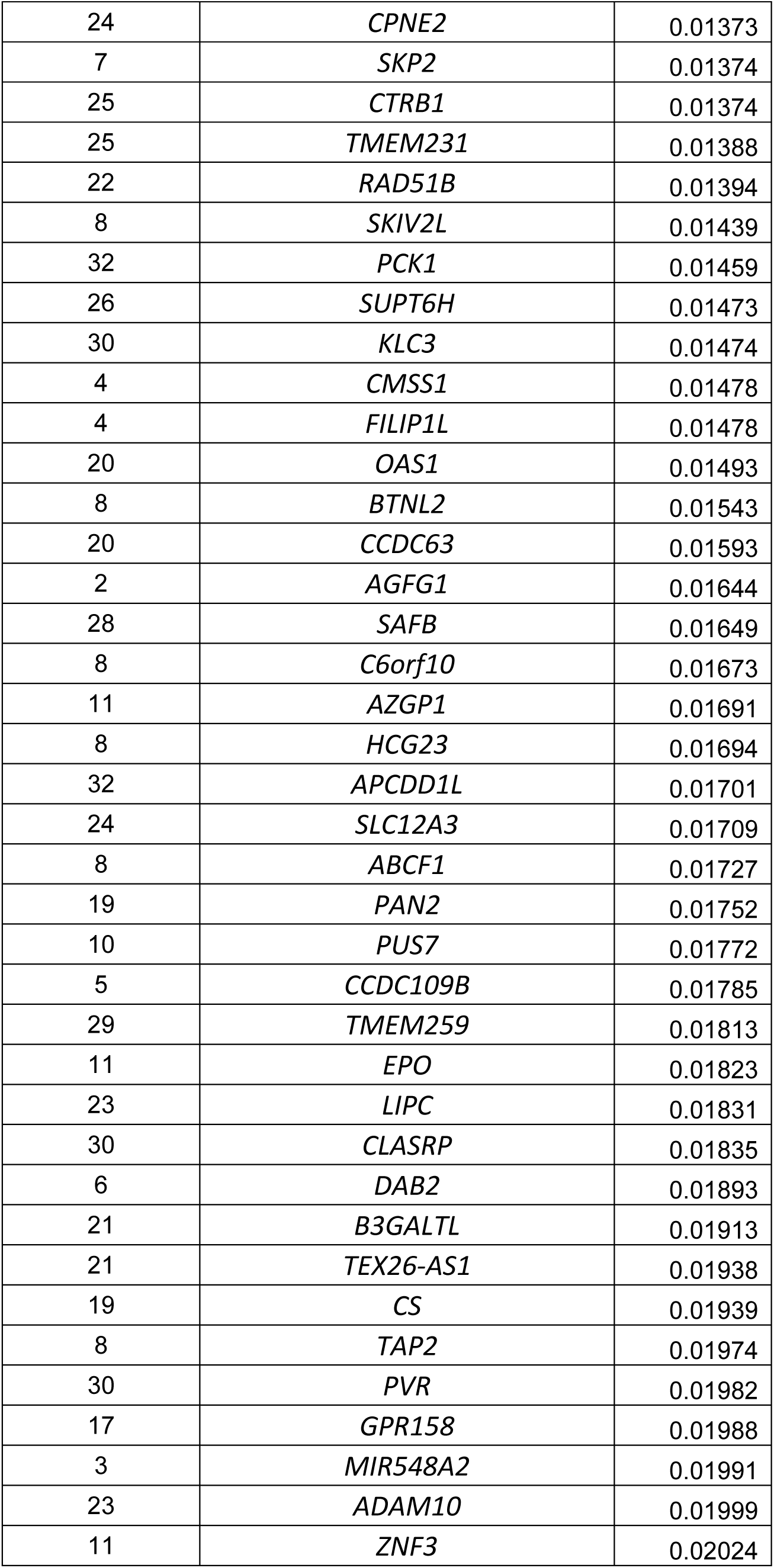
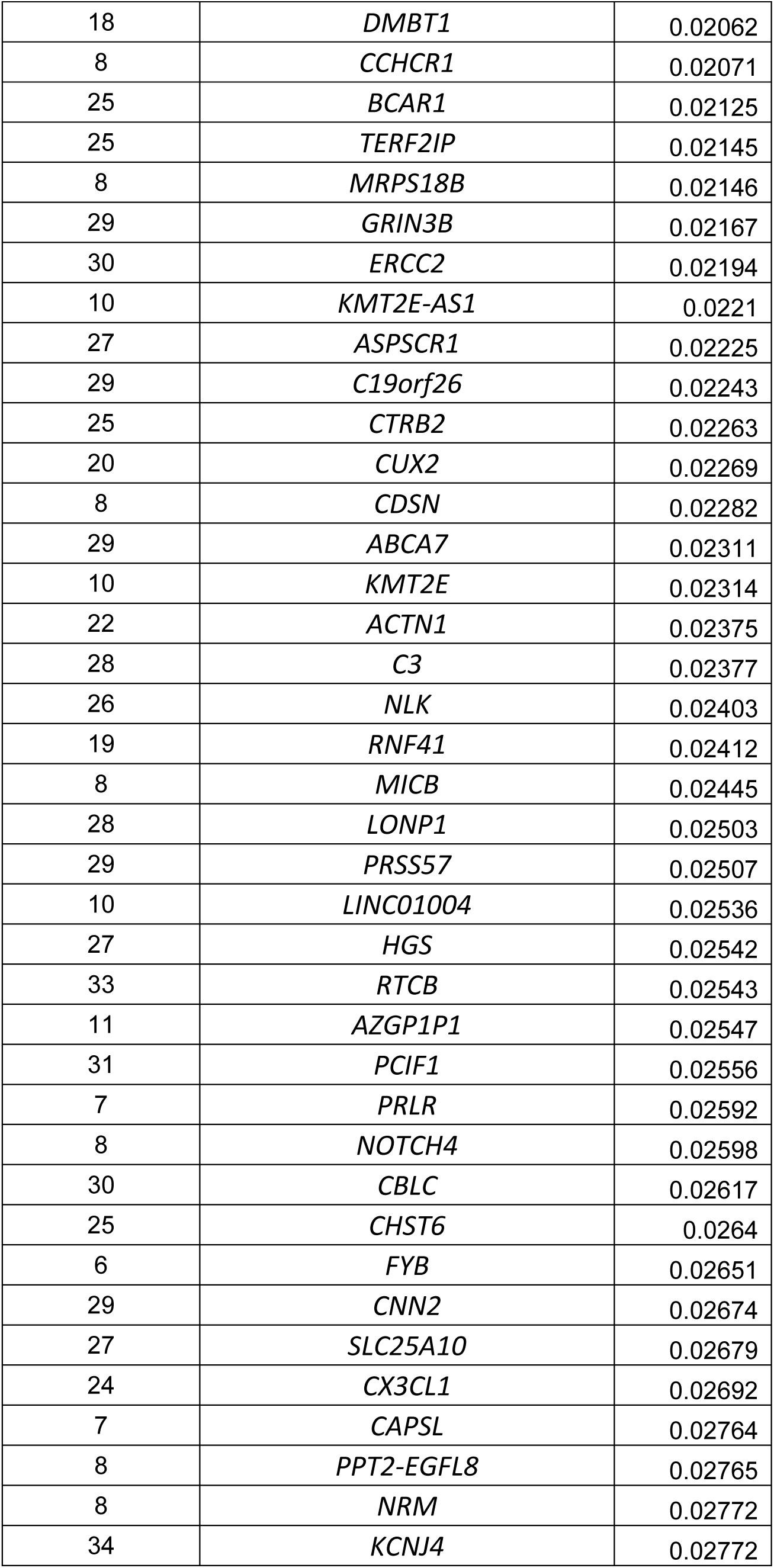
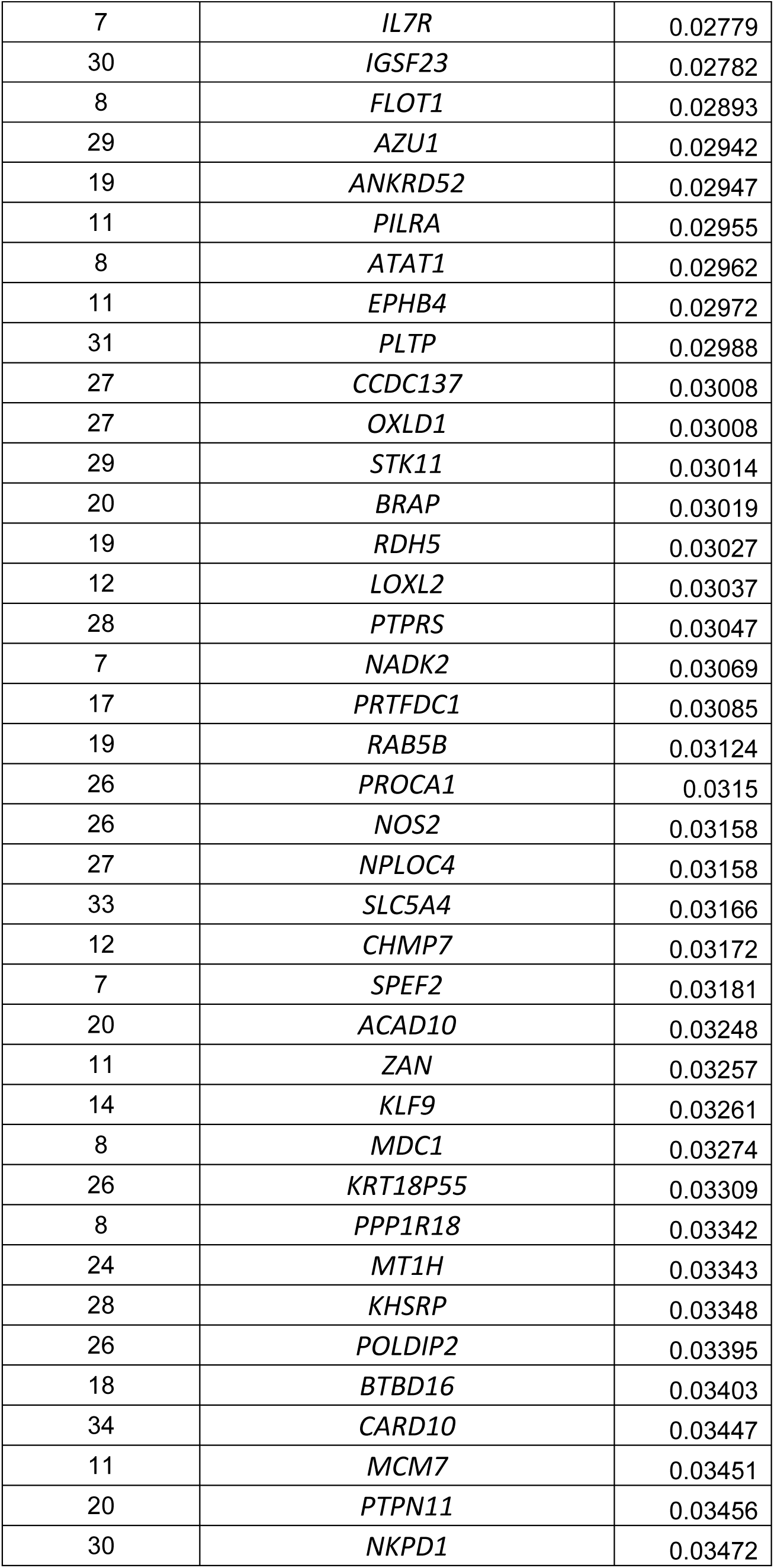
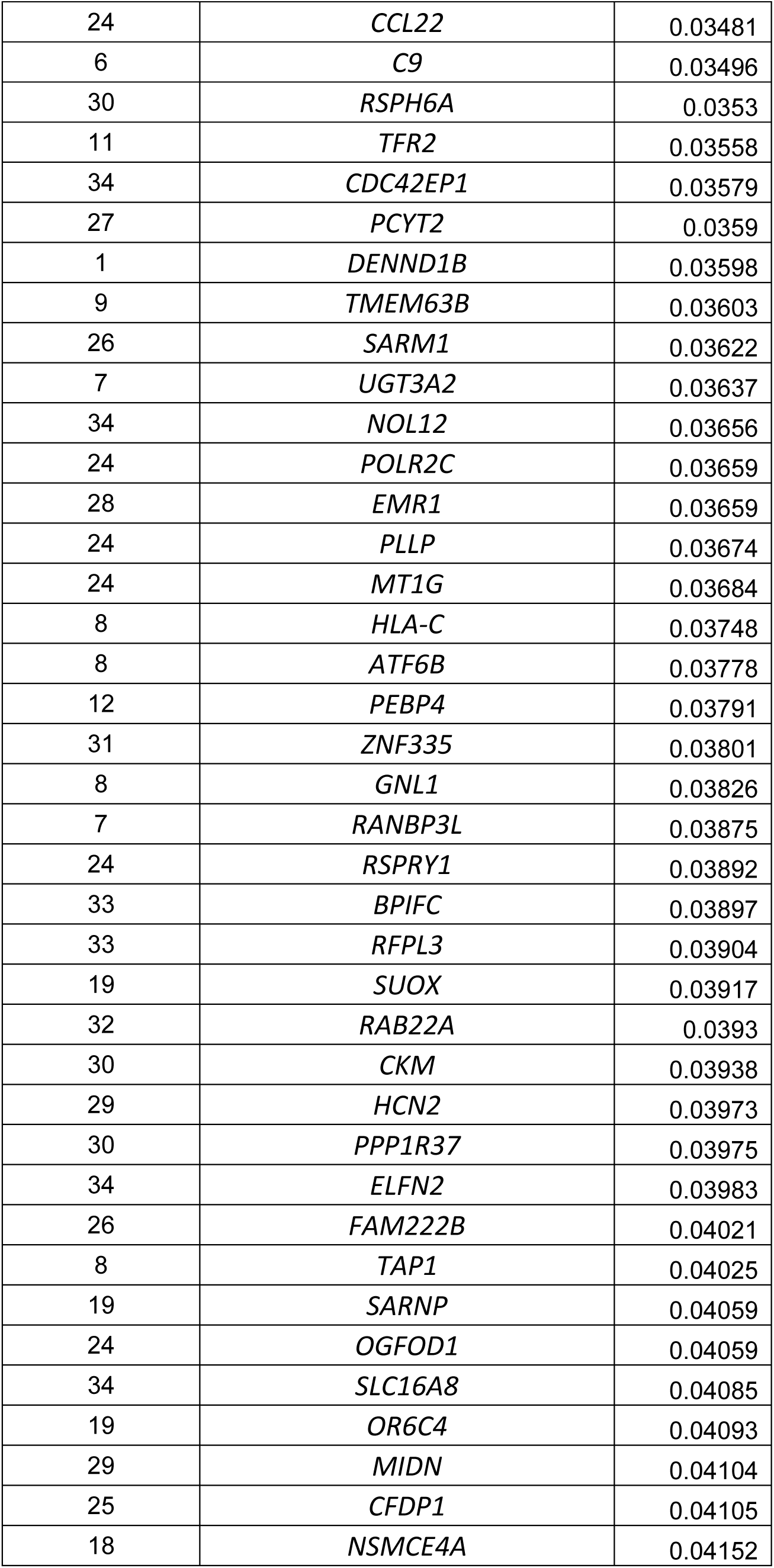
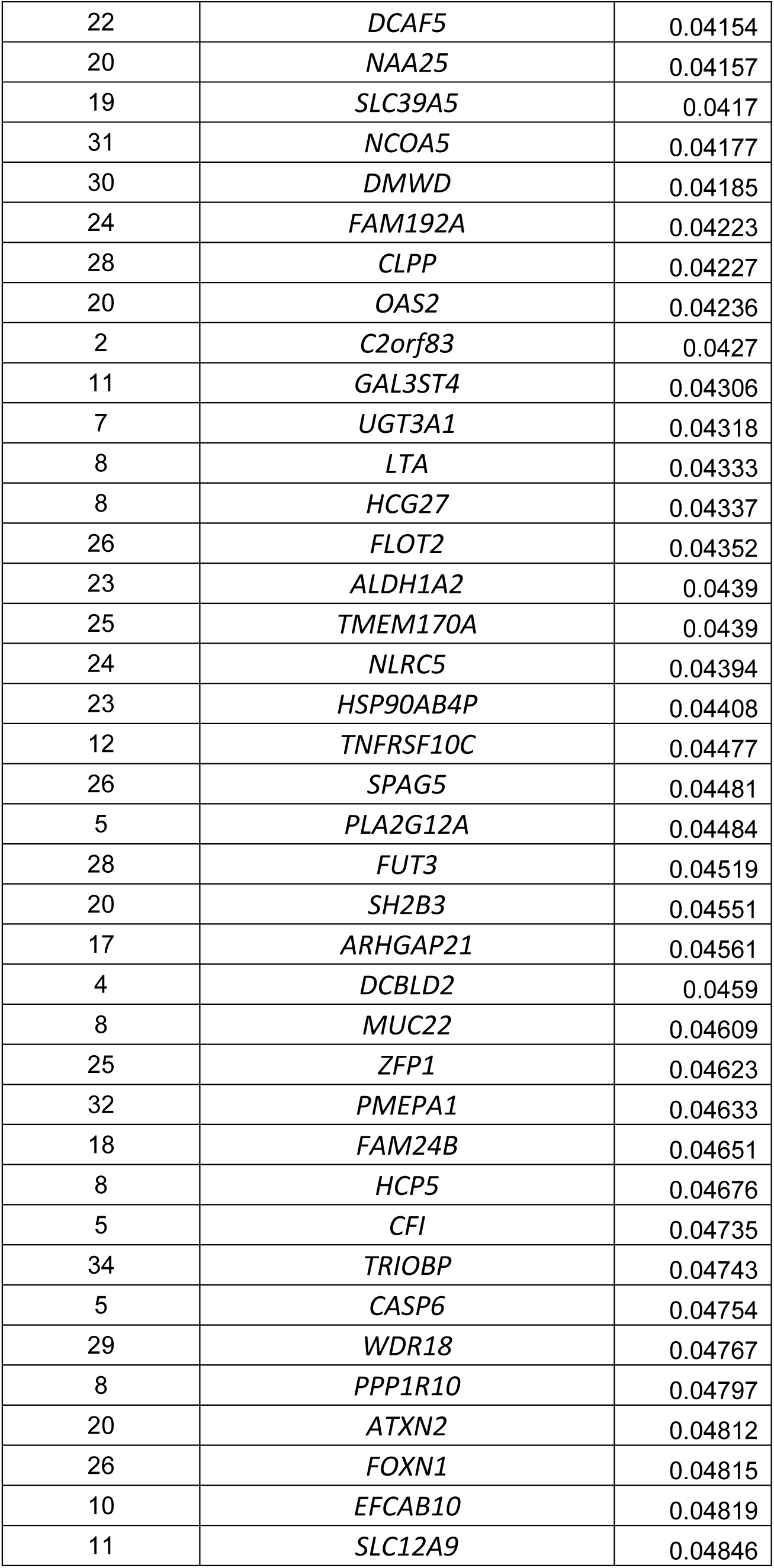
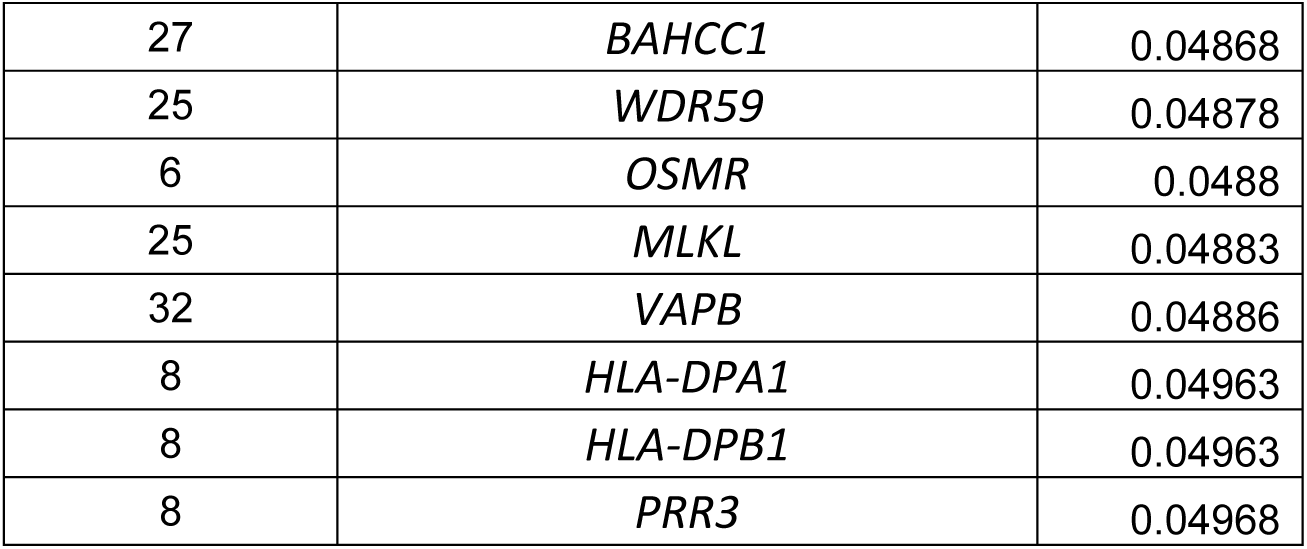
Association statistics of variants in known AMD risk loci that were nominally associated with AMD in the Israeli discovery set (P<0.05, 31/34 loci).

**Supplementary Table 4.**
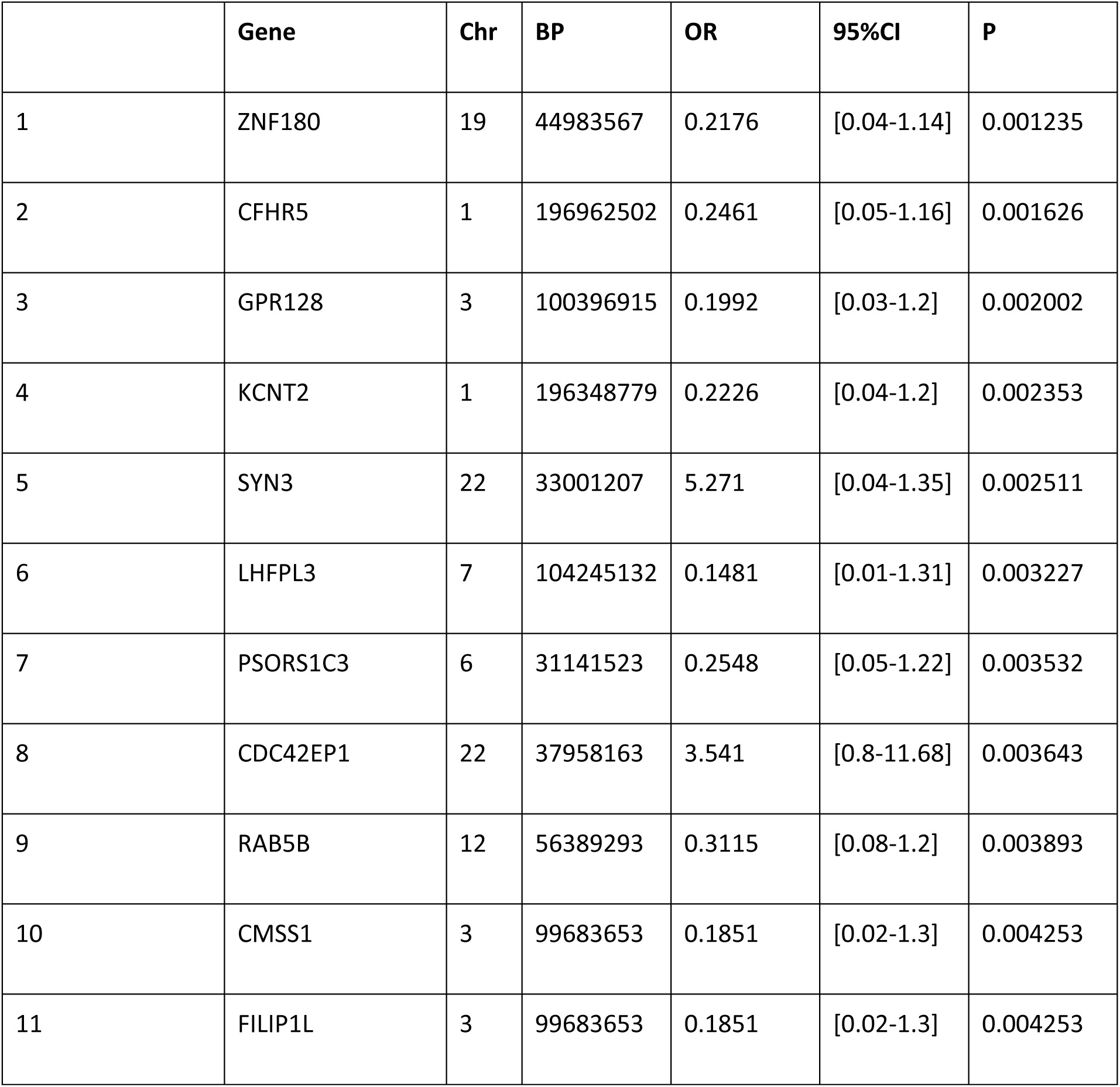
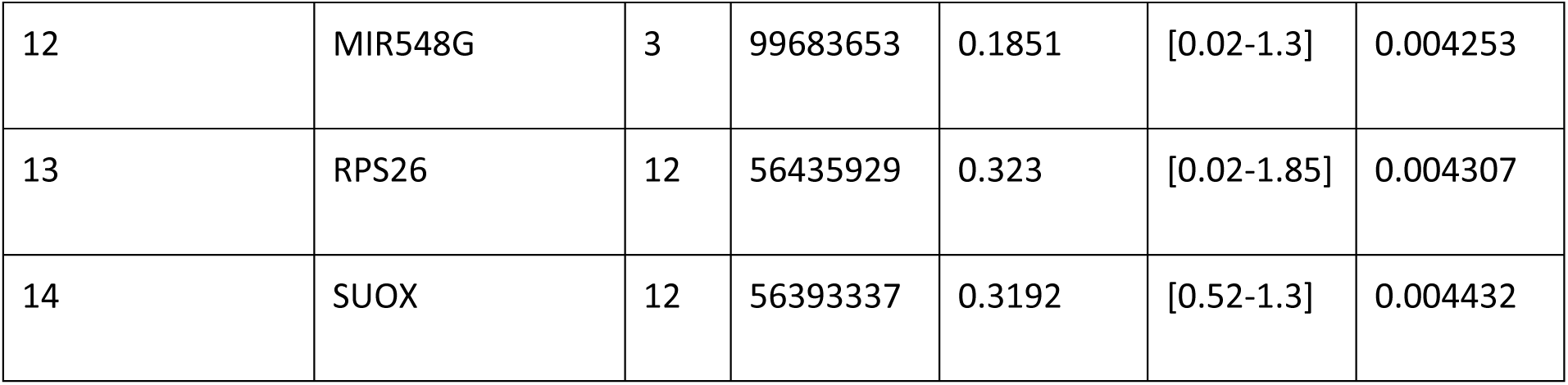
Association statistics of variants in known AMD risk loci in the Ashkenazi subpopulation (P<1x10^-4^). The data is reported as in Supplementary Table 3.

|  | **Gene** | **Chr** | **BP** | **OR** | **95%CI** | **P** |
| --- | --- | --- | --- | --- | --- | --- |
| 1 | ZNF180 | 19 | 44983567 | 0.2176 | [0.04-1.14] | 0.001235 |
| 2 | CFHR5 | 1 | 196962502 | 0.2461 | [0.05-1.16] | 0.001626 |
| 3 | GPR128 | 3 | 100396915 | 0.1992 | [0.03-1.2] | 0.002002 |
| 4 | KCNT2 | 1 | 196348779 | 0.2226 | [0.04-1.2] | 0.002353 |
| 5 | SYN3 | 22 | 33001207 | 5.271 | [0.04-1.35] | 0.002511 |
| 6 | LHFPL3 | 7 | 104245132 | 0.1481 | [0.01-1.31] | 0.003227 |
| 7 | PSORS1C3 | 6 | 31141523 | 0.2548 | [0.05-1.22] | 0.003532 |
| 8 | CDC42EP1 | 22 | 37958163 | 3.541 | [0.8-11.68] | 0.003643 |
| 9 | RAB5B | 12 | 56389293 | 0.3115 | [0.08-1.2] | 0.003893 |
| 10 | CMSS1 | 3 | 99683653 | 0.1851 | [0.02-1.3] | 0.004253 |
| 11 | FILIP1L | 3 | 99683653 | 0.1851 | [0.02-1.3] | 0.004253 |
| 12 | MIR548G | 3 | 99683653 | 0.1851 | [0.02-1.3] | 0.004253 |
| 13 | RPS26 | 12 | 56435929 | 0.323 | [0.02-1.85] | 0.004307 |
| 14 | SUOX | 12 | 56393337 | 0.3192 | [0.52-1.3] | 0.004432 |

**Supplementary Table 5.** Association statistics of variants in known AMD risk loci in the Arab subpopulation (P<1x10^-4^). The data is reported as in Supplementary Table 3.

|  | Gene | Chr | BP | OR | 95%CI | P |
| --- | --- | --- | --- | --- | --- | --- |
| 1 | HTRA1 | 10 | 124221270 | 2.221 | [1.08-4.58] | 2.914E-06 |
| 2 | ARMS2 | 10 | 124215211 | 2.23 | [1.08-4.6] | 3.062E-06 |
| 3 | CFH | 1 | 196673430 | 1.984 | [1.02-3.84] | 0.00003481 |
| 4 | CFHR2 | 1 | 196927791 | 1.931 | [1.01-3.68] | 0.00006749 |
| 5 | ZNF557 | 19 | 7083629 | 1.823 | [0.98-3.4] | 0.0003984 |
| 6 | SRPK2 | 7 | 104877373 | 0.5753 | [0.32-1.03] | 0.0005616 |
| 7 | TOMM40 | 19 | 45396219 | -3.424 | [0.25-1.04] | 0.0006182 |
| 8 | CFHR5 | 1 | 196978615 | 0.582 | [0.32-1.03] | 0.0007713 |
| 9 | HLA-B | 6 | 31324864 | 0.3546 | [0.11-1.08] | 0.0009341 |
| 10 | CFHR4 | 1 | 196870299 | 0.3862 | [0.14-1.07] | 0.0009646 |
| 11 | PLEKHA1 | 10 | 124139393 | 1.695 | [0.95-3] | 0.0009873 |

| dbSNP Identifier | HGVS nomenclature | Chr:BP |
| --- | --- | --- |
| rs1195500:G>A,C,T | NC_000015.10:g.29394842G>A,<br>NC_000015.10:g.29394842G>C,<br>NC_000015.10:g.29394842G>T | Chr15:29687047 (GRCh37) |
| rs116928937:C>A | NC_000015.10:g.65385107C>A | Chr15:65677446 (GRCh37) |
| rs142491581:C>G | NC_000004.12:g.127714391C>G | Chr4:128635546 (GRCh37) |
| rs6449549:C>T | NC_000005.10:g.61734604C>T | Chr5:61030432 (GRCh37) |
| rs4235321:G>A | NC_000004.12:g.24954305G>A | Chr4:24955928 (GRCh37) |
| rs41592:G>A | NC_000007.14:g.83012543G>A | Chr7:82641860 (GRCh37) |
| rs12701455:A>G,T | NC_000007.14:g.1015773A>G,N<br>C_000007.14:g.1015773A>T | Chr7:1055409 (GRCh37) |
| rs11689931:T>G | NC_000002.12:g.205576255T>G | Chr2:206440979 (GRCh37) |
| rs1506825:A>C,T | NC_000016.10:g.76449122A>C,N<br>C_000016.10:g.76449122A>T | Chr16:76483019 (GRCh37) |

| Gene | dbSNP TopVariant | HGVS | TopVariant:Chr:BP<br>(GRCh37) |
| --- | --- | --- | --- |
| CFH | rs3766405:C>A,T | NC_000001.11:g.196726031C><br>A,NC_000001.11:g.196726031<br>C>T | Chr1:196695161 |
| ARMS2 | rs10490924:G>C,T | NC_000010.11:g.122454932G><br>C,NC_000010.11:g.122454932<br>G>T | Chr10:124214448 |
| HTRA1 | rs1049331:C>T | NC_000010.11:g.122461753C><br>T | Chr10:124221270 |
| CFHR5 | rs10922153:T>G | NC_000001.11:g.197009484T><br>G | Chr1:196978615 |
| CFHR2 | rs2026547:G>A | NC_000001.11:g.196958661G><br>A | Chr1:196927791 |
| CFHR4 | rs34833349:A>G | NC_000001.11:g.196901169A><br>G | Chr1:196870299 |
| KCNT2 | rs10922068:T>A,C | NC_000001.11:g.196437585T>A,NC_000001.11:g.196437585T>C | Chr1:196406715 |
| F13B | rs10754210:G>A | NC_000001.11:g.197042981G>A | Chr1:197012111 |
| SYN3 | rs5754187:C>T | NC_000022.11:g.32651611C>T | Chr22:33047598 |
| ASPM | rs6676084:C>T | NC_000001.11:g.197124900C>T | Chr1:197094030 |
| ZBTB41 | rs4350226:G>A,C,T | NC_000001.11:g.197163247G>A,NC_000001.11:g.197163247G>C,NC_000001.11:g.197163247G>T | Chr1:197132378 |
| PLEKHA1 | rs2421017:A>G | NC_000010.11:g.122388651A>G | Chr10:124148167 |
| VPS29 | rs184629901:T>C | NC_000012.12:g.110497463T>C | Chr12:110935268 |
| CRB1 | rs12737179:T>C,G | NC_000001.11:g.197230303T>C,NC_000001.11:g.197230303T>G | Chr1:197199434 |
| PPTC7 | rs56159960:T>C | NC_000012.12:g.110556262T>C | Chr12:110994068 |
| ZNF557 | rs966591:A>C,G | NC_000019.10:g.7083618A>C,NC_000019.10:g.7083618A>G | Chr19: 7083629 |
| HVCN1 | rs73191857:C>T | NC_000012.12:g.110661916C>T | Chr12: 111099721 |
| MICALL1 | rs9607501:G>A,C,T | NC_000022.11:g.37922890G>A,NC_000022.11:g.37922890G>C,NC_000022.11:g.37922890G>T | Chr22: 38318897 |
| SMC5 | rs66524845:A>G | NC_000009.12:g.70305808A>G | Chr9: 72920724 |
| PPP1CC | rs1973505:G>A | NC_000012.12:g.110722198G>A | Chr12: 111160003 |
| TCTN1 | rs7953794:A>G | NC_000012.12:g.110643392A>G | Chr12: 111081197 |
| HLA-B | rs151341076:G>A,C,T | NC_000006.12:g.31357087G>A,NC_000006.12:g.31357087G>C,NC_000006.12:g.31357087G>T | Chr6:31324864 |
| CFHR5 | rs7547265:G>C,T | NC_000001.11:g.196993372G>C,NC_000001.11:g.196993372G>T | Chr1:196962502 |
| GPR128 | rs7629279:G>T | NC_000003.12:g.100678070G>T | Chr3:100396915 |
| KCNT2 | rs7527415:C>T | NC_000001.11:g.196379649C>T | Chr1:196348779 |
| LHFPL3 | rs17139096:A>G,T | NC_000007.14:g.104604684A>G,NC_000007.14:g.104604684A>T | Chr7:104245132 |
| PSORS1C<br>3 | rs887468:C>T | NC_000006.12:g.31173746C>T | Chr6:31141523 |
| CDC42EP<br>1 | rs2235335:G>A,C | NC_000022.11:g.37562156G>A,NC_000022.11:g.37562156G>C | Chr22:37958163 |
| RAB5B | rs705700:T>A,C | NC_000012.12:g.55995509T>A,NC_000012.12:g.55995509T>C | Chr12:56389293 |
| FILIP1L | rs73138610:C>A,G | NC_000003.12:g.99964809C>A,NC_000003.12:g.99964809C>G | Chr3:99683653 |
| RPS26 | rs1131017:C>A,G,T | NC_000012.12:g.56042145C>A,NC_000012.12:g.56042145C>G,NC_000012.12:g.56042145C>T | Chr12:56435929 |
| SUOX | rs1081975:C>A,G | NC_000012.12:g.55999553C>A,NC_000012.12:g.55999553C>G | Chr12:56393337 |
| HTRA1 | rs1049331:C>T | NC_000010.11:g.122461753C>T | Chr10:124221270 |
| ARMS2 | rs36212733:T>A,C,G | NC_000010.11:g.122455695T>A,NC_000010.11:g.122455695T>C,NC_000010.11:g.122455695T>G | Chr10:124215211 |
| CFH | rs9970075:T>A,G | NC_000001.11:g.196704299T>A,NC_000001.11:g.196704299T>G | Chr1:196673430 |
| CFHR2 | rs2026547:G>A | NC_000001.11:g.196958661G>A | Chr1:196927791 |
| ZNF557 | rs966591:A>C,G | NC_000019.10:g.7083618A>C,<br>NC_000019.10:g.7083618A>G | Chr19:7083629 |
| SRPK2 | rs6950104:G>A,C | NC_000007.14:g.105236926G><br>A,NC_000007.14:g.105236926<br>G>C | Chr7:104877373 |
| TOMM4<br>0 | rs157582:C>T | NC_000019.10:g.44892961C>T | Chr19:45396219 |
| CFHR5 | rs10922153:T>G | NC_000001.11:g.197009484T><br>G | Chr1:196978615 |
| HLA-B | rs151341076:G>A,C,T | NC_000006.12:g.31357087G><br>A,NC_000006.12:g.31357087G<br>>C,<br>NC_000006.12:g.31357087G>T | Chr6:31324864 |
| CFHR4 | rs34833349:A>G | NC_000001.11:g.196901169A><br>G | Chr1:196870299 |
| PLEKHA1 | rs11200594:C>G,T | NC_000010.11:g.122379876C><br>G,NC_000010.11:g.122379876<br>C>T | Chr10:124139393 |

## References

1. DeAngelis MM, Owen LA, Morrison MA, et al. Genetics of age-related macular degeneration (AMD). Hum Mol Genet. 2017;26(R1):R45–R50. doi:10.1093/hmg/ddx228

2. Fritsche LGLG, Igl W, Bailey JNCJNC, et al. A large genome-wide association study of age-related macular degeneration highlights contributions of rare and common variants. Nat Genet. 2016;48(2):134–143. doi:10.1038/ng.3448

3. Haines JL, Hauser MA, Schmidt S, et al. Complement factor H variant increases the risk of age-related macular degeneration. Science (80-). 2005;308(5720):419–421. http://www.ncbi.nlm.nih.gov/entrez/query.fcgi?cmd=Retrieve&db=PubMed&dopt=Citation&list_uids=15761120.

4. Klein RJ, Zeiss C, Chew EY, et al. Complement factor H polymorphism in age-related macular degeneration. Science (80-). 2005;308(5720):385–389. http://www.ncbi.nlm.nih.gov/entrez/query.fcgi?cmd=Retrieve&db=PubMed&dopt=Citation&list_uids=15761122.

5. Behar DM, Yunusbayev B, Metspalu M, et al. The genome-wide structure of the Jewish people. Nature. 2010;466(7303):238–242. doi:10.1038/nature09103

6. Carmi S, Hui KY, Kochav E, et al. Sequencing an Ashkenazi reference panel supports population-targeted personal genomics and illuminates Jewish and European origins. Nat Commun. 2014;5:4835. doi:10.1038/ncomms5835

7. Waldman S, Backenroth D, Harney É, et al. Genome-wide data from medieval German Jews show that the Ashkenazi founder event pre-dated the 14(th) century. Cell. 2022;185(25):4703–4716.e16. doi:10.1016/j.cell.2022.11.002

8. Agranat-Tamir L, Waldman S, Martin MAS, et al. The Genomic History of the Bronze Age Southern Levant. Cell. 2020;181(5):1146–1157.e11. doi:10.1016/j.cell.2020.04.024

9. Granot-Hershkovitz E, Karasik D, Friedlander Y, et al. A study of Kibbutzim in Israel reveals risk factors for cardiometabolic traits and subtle population structure. Eur J Hum Genet. 2018;26(12):1848–1858. doi:10.1038/s41431-018-0230-3

10. Zidan J, Ben-Avraham D, Carmi S, Maray T, Friedman E, Atzmon G. Genotyping of geographically diverse Druze trios reveals substructure and a recent bottleneck. Eur J Hum Genet. 2014;8(February):1093–1099. doi:10.1038/ejhg.2014.218

11. Zeggini E. Using genetically isolated populations to understand the genomic basis of disease. Genome Med. 2014;6(10):83. doi:10.1186/s13073-014-0083-5

12. Zelinger L, Banin E, Obolensky A, et al. A missense mutation in DHDDS, encoding dehydrodolichyl diphosphate synthase, is associated with autosomal-recessive retinitis pigmentosa in ashkenazi jews. Am J Hum Genet. 2011;88(2):207–215. doi:10.1016/j.ajhg.2011.01.002

13. Zlotogora J, Chemke J. Medical genetics in Israel. Eur J Hum Genet. 1995;3(3):147–154. http://www.ncbi.nlm.nih.gov/entrez/query.fcgi?cmd=Retrieve&db=PubMed&dopt=Citation&list_uids=7583040.

14. Beryozkin A, Shevah E, Kimchi A, et al. Whole Exome Sequencing Reveals Mutations in Known Retinal Disease Genes in 33 out of 68 Israeli Families with Inherited Retinopathies. Sci Rep. 2015;5. doi:10.1038/srep13187

15. Chowers I, Cohen Y, Goldenberg-Cohen N, et al. Association of complement factor H Y402H polymorphism with phenotype of neovascular age related macular degeneration in Israel. Mol Vis. 2008;14:1829–1834. http://www.ncbi.nlm.nih.gov/entrez/query.fcgi?cmd=Retrieve&db=PubMed&dopt=Citation&list_uids=18852870.

16. Chowers I, Meir T, Lederman M, et al. Sequence variants in HTRA1 and LOC387715/ARMS2 and phenotype and response to photodynamic therapy in neovascular age-related macular degeneration in populations from Israel. Mol Vis. 2008;14:2263–2271. http://www.ncbi.nlm.nih.gov/entrez/query.fcgi?cmd=Retrieve&db=PubMed&dopt=Citation&list_uids=19065273.

17. Asleh SAA, Lederman M, Weinstein O, et al. Lack of association between the C2 allele of transferrin and age-related macular degeneration in the Israeli population. Ophthalmic Genet. 2009;30(4):161–164. doi:10.3109/13816810903147998 [pii] 10.3109/13816810903147998

18. Babb de Villiers C, Kroese M, Moorthie S. Understanding polygenic models, their development and the potential application of polygenic scores in healthcare. J Med Genet. 2020;57(11):725–732. doi:10.1136/jmedgenet-2019-106763

19. Torkamani A, Wineinger NE, Topol EJ. The personal and clinical utility of polygenic risk scores. Nat Rev Genet. 2018. doi:10.1038/s41576-018-0018-x

20. Khera A V., Chaffin M, Aragam KG, et al. Genome-wide polygenic scores for common diseases identify individuals with risk equivalent to monogenic mutations. Nat Genet. 2018. doi:10.1038/s41588-018-0183-z

21. Heesterbeek TJ, de Jong EK, Acar IE, et al. Genetic risk score has added value over initial clinical grading stage in predicting disease progression in age-related macular degeneration. Sci Rep. 2019;9(1):6611. doi:10.1038/s41598-019-43144-3

22. Colijn JM, Meester-Smoor M, Verzijden T, et al. Genetic Risk, Lifestyle, and Age-Related Macular Degeneration in Europe: The EYE-RISK Consortium. Ophthalmology. 2021;128(7):1039–1049. doi:10.1016/j.ophtha.2020.11.024

23. Martin AR, Kanai M, Kamatani Y, Okada Y, Neale BM, Daly MJ. Clinical use of current polygenic risk scores may exacerbate health disparities. Nat Genet. 2019;51(4):584–591. doi:10.1038/s41588-019-0379-x

24. Duncan L, Shen H, Gelaye B, et al. Analysis of polygenic risk score usage and performance in diverse human populations. Nat Commun. 2019;10(1):3328. doi:10.1038/s41467-019-11112-0

25. Adzhubei I, Jordan DM, Sunyaev SR. Predicting functional effect of human missense mutations using PolyPhen-2. Curr Protoc Hum Genet. 2013;(SUPPL.76). doi:10.1002/0471142905.hg0720s76

26. Vilhjálmsson BJ, Yang J, Finucane HK, et al. Modeling Linkage Disequilibrium Increases Accuracy of Polygenic Risk Scores. Am J Hum Genet. 2015;97(4):576–592. doi:10.1016/j.ajhg.2015.09.001

27. Privé F, Arbel J, Vilhjálmsson BJ. LDpred2: better, faster, stronger. Bioinformatics. 2020;36(22-23):5424–5431. doi:10.1093/bioinformatics/btaa1029

28. Thomas M, Sakoda LC, Hoffmeister M, et al. Genome-wide Modeling of Polygenic Risk Score in Colorectal Cancer Risk. Am J Hum Genet. 2020;107(3):432–444. doi:10.1016/j.ajhg.2020.07.006

29. Vaura F, Kauko A, Suvila K, et al. Polygenic Risk Scores Predict Hypertension Onset and Cardiovascular Risk. Hypertens (Dallas, Tex 1979). 2021;77(4):1119–1127. doi:10.1161/HYPERTENSIONAHA.120.16471

30. Qassim A, Souzeau E, Hollitt G, Hassall MM, Siggs OM, Craig JE. Risk Stratification and Clinical Utility of Polygenic Risk Scores in Ophthalmology. Transl Vis Sci Technol. 2021;10(6):14. doi:10.1167/tvst.10.6.14

31. Fritsche LG, Igl W, Bailey JNC, et al. A large genome-wide association study of age-related macular degeneration highlights contributions of rare and common variants. Nat Genet. 2016;48(2):134–143. doi:10.1038/ng.3448

32. Gettler K, Levantovsky R, Moscati A, et al. Common and Rare Variant Prediction and Penetrance of IBD in a Large, Multi-ethnic, Health System-based Biobank Cohort. Gastroenterology. 2021;160(5):1546–1557. doi:10.1053/j.gastro.2020.12.034

33. Belbin GM, Cullina S, Wenric S, et al. Toward a fine-scale population health monitoring system. Cell. 2021;184(8):2068–2083.e11. doi:10.1016/j.cell.2021.03.034

34. Fahed AC, Aragam KG, Hindy G, et al. Transethnic Transferability of a Genome-Wide Polygenic Score for Coronary Artery Disease. Circ Genomic Precis Med. 2021;14(1):e003092. doi:10.1161/CIRCGEN.120.003092

35. Privé F, Aschard H, Carmi S, et al. Portability of 245 polygenic scores when derived from the UK Biobank and applied to 9 ancestry groups from the same cohort. Am J Hum Genet. 2022;109(1):12–23. doi:10.1016/j.ajhg.2021.11.008

36. Cai M, Xiao J, Zhang S, et al. A unified framework for cross-population trait prediction by leveraging the genetic correlation of polygenic traits. Am J Hum Genet. 2021;108(4):632–655. doi:10.1016/j.ajhg.2021.03.002

37. Kachuri L, Chatterjee N, Hirbo J, et al. Principles and methods for transferring polygenic risk scores across global populations. Nat Rev Genet. August 2023. doi:10.1038/s41576-023-00637-2

38. Lorés-Motta L, Riaz M, Grunin M, et al. Association of genetic variants with response to anti-vascular endothelial growth factor therapy in age-related macular degeneration. JAMA Ophthalmol. 2018. doi:10.1001/jamaophthalmol.2018.2019

39. Age-Related Eye Disease Study Research G. The Age-Related Eye Disease Study (AREDS): design implications. AREDS report no. 1. Control Clin Trials. 1999;20(6):573–600. http://www.ncbi.nlm.nih.gov/pubmed/10588299.

40. The 1000 Genomes Project Consortium. A global reference for human genetic variation. Nature. 2015;526(7571):68–74. doi:10.1038/nature15393

41. Lencz T, Yu J, Palmer C, et al. High-depth whole genome sequencing of an Ashkenazi Jewish reference panel: enhancing sensitivity, accuracy, and imputation. Hum Genet. 2018;137(4):343–355. doi:10.1007/s00439-018-1886-z

42. Delaneau O, Marchini J, Zagury J-F. A linear complexity phasing method for thousands of genomes. Nat Methods. 2012;9(2):179–181. doi:10.1038/nmeth.1785

43. Delaneau O, Zagury J-F, Marchini J. Improved whole-chromosome phasing for disease and population genetic studies. Nat Methods. 2013;10(1):5–6. doi:10.1038/nmeth.2307

44. van Leeuwen EM, Kanterakis A, Deelen P, et al. Population-specific genotype imputations using minimac or IMPUTE2. Nat Protoc. 2015;10(9):1285–1296. doi:10.1038/nprot.2015.077

45. Grunin M, Beykin G, Rahmani E, et al. Association of a variant in VWA3A with response to anti-vascular endothelial growth factor treatment in neovascular AMD. Investig Ophthalmol Vis Sci. 2020. doi:10.1167/iovs.61.2.48

46. Anderson CA, Pettersson FHFHFH, Clarke GMGMGM, Cardon LR, Morris AP, Zondervan KT. Data quality control in genetic case-control association studies. Nat Protoc. 2010;5(9):1564–1573. doi:10.1038/nprot.2010.116

47. Chang CC, Chow CC, Tellier LC a. M, Vattikuti S, Purcell SM, Lee JJ. Second-generation PLINK: rising to the challenge of larger and richer datasets. 2014:1–22. http://arxiv.org/abs/1410.4803v1.

48. Purcell S, Neale B, Todd-Brown K, et al. PLINK: A tool set for whole-genome association and population-based linkage analyses. Am J Hum Genet. 2007;81(3):559–575. doi:10.1086/519795

49. Yang J, Lee SH, Goddard ME, Visscher PM. GCTA: A tool for genome-wide complex trait analysis. Am J Hum Genet. 2011;88(1):76–82. doi:10.1016/j.ajhg.2010.11.011

50. Privé F, Vilhjálmsson BJ, Aschard H, Blum MGB. Making the Most of Clumping and Thresholding for Polygenic Scores. Am J Hum Genet. 2019;105(6):1213–1221. 10.1016/j.ajhg.2019.11.001

51. Choi SW, Mak TS-H, O’Reilly PF. Tutorial: a guide to performing polygenic risk score analyses. Nat Protoc. 2020;15(9):2759–2772. doi:10.1038/s41596-020-0353-1

